# Altered Auditory Network Differentiation in Multiple Sclerosis: A BROAD-NESS MEG Study

**DOI:** 10.64898/2026.11.25.26368990

**Authors:** Francesco Carlomagno, Leonardo Bonetti, Mattia Rosso, Elvira Brattico, Paolo Taurisano, Madia Marika Biasi, Giusy Tancredi, Alessia Manni, Antonio Iaffaldano, Giulia Paparella, Daphne Gasparre, Marina de Tommaso, Vitoantonio Bevilacqua, Damiano Paolicelli

## Abstract

The auditory P300 is widely used at the sensor level to study cognitive dysfunction in multiple sclerosis (MS), but its brain generators have rarely been investigated. The three-stimulus oddball separates task relevance from stimulus rarity, and source-level network analysis can show where and when this distinction is altered. We recorded magnetoencephalography (MEG) in 20 MS patients and 15 healthy controls during a three-stimulus auditory oddball with standard, target and non-target tones. Broadband networks were estimated from beamformer source activity with BROAD-NESS, a principal component analysis of group-averaged sources. Groups were compared with cluster-based permutation tests corrected across networks. Four networks explained 94% of the variance. Sensory (N1m) and P3 responses and task performance did not differ between groups. In a bilateral perisylvian and sensorimotor network, the difference between target and non-target responses was smaller in patients than in controls at about 370–500 ms (cluster p = 0.020, corrected across networks; Hedges’ g = −1.11 and Bayes factor 16.1 in the localizer window). A conventional source-by-source analysis of the same contrast found no cluster surviving correction. The effect was robust to leave-one-out analysis, an alternative network estimation and excluding participants with clinical or recording characteristics, but did not survive correction across all three contrasts (p = 0.108). Network measures were not related to discrimination performance or fatigue, and no exploratory correlation with clinical or cognitive measures survived correction. In MS, task-related source-level networks were largely preserved, but one network differentiated less between task-relevant and task-irrelevant deviants, without a behavioural deficit.

**Highlights:**

- Four broadband MEG source networks described a three-stimulus auditory oddball
- N1m and P3 network responses did not differ between MS patients and controls
- An auditory network separated targets from non-targets less in MS patients
- The network difference occurred without differences in task performance
- Network measures showed no reliable relation to fatigue or cognition

## 1. Introduction

Cognitive impairment is common in multiple sclerosis (MS) and most often affects information processing speed and attention (Chiaravalloti and DeLuca, 2008). Because it can be present early and is not well captured by disability scales, there is interest in neurophysiological markers that index the underlying cognitive processes. The auditory oddball paradigm and its P300 response have been used most widely for this purpose. The auditory pathways are relatively spared in MS (Sanders et al., 1986), so that late cortical responses can be studied with limited confounding from afferent dysfunction.

Event-related potential studies have reported prolonged P300 latency and, less consistently, reduced P300 amplitude in patients with MS, with associations with neuropsychological performance, disability and lesion load (Magnano et al., 2006; Whelan et al., 2010; Sundgren et al., 2015). Changes have also been described in early relapsing-remitting MS and in clinically isolated syndrome (Kocer et al., 2008; Pokryszko-Dragan et al., 2016). Paolicelli et al. (2021) recorded simultaneous MEG and high-density EEG during a two-stimulus auditory oddball in MS patients and healthy controls and analysed the responses at the sensor level. P300 latency was slightly but not significantly longer in patients, P300 amplitude and scalp topography were similar in the two groups, and time–frequency analysis showed lower induced MEG power to standard tones in patients; P300 amplitude correlated inversely with fatigue in the pooled sample of patients and controls. Sensor-level measures of evoked brain responses may therefore be relatively insensitive to alterations in MS, and the authors pointed to the cortical generators of the response as the next step. Source-level studies of the P3 in MS remain few. A high-density EEG study localized altered frontoparietal P3 generators during visual working memory in newly diagnosed patients (Arrabal et al., 2026), and an MEG study of an n-back task found a frontoparietal network active 300–600 ms after the stimulus, resembling the P300, alongside a reduced early prefrontal network in patients (Rossi et al., 2024). To our knowledge, the generators of auditory deviance responses, and how they separate task-relevant from task-irrelevant sounds, have not been examined in MS.

The classic two-stimulus oddball cannot separate the processing of a rare stimulus from that of a task-relevant one, because the rare stimulus is also the target. The three-stimulus oddball adds a rare non-target stimulus that must be ignored (Squires et al., 1975). Rare non-targets elicit a fronto-central P3a, linked to the orienting of attention, whereas targets elicit a parietal P3b, linked to task-relevant stimulus evaluation (Polich, 2007). Comparing target and non-target responses therefore isolates task relevance at matched probability. P3a and P3b amplitudes have been related to PASAT performance in progressive MS (Kiiski et al., 2011), but how these processes are organized at the level of brain networks in MS is unknown.

Source reconstruction of MEG data can address this question, but analyses restricted to a priori regions or time windows can miss distributed effects and invite circular selection. BROAD-NESS (Bonetti et al., 2025a) estimates broadband brain networks directly from source-reconstructed evoked responses. Principal component analysis of the group-averaged source activity yields a small number of spatial patterns, and the time course of each pattern can be compared across conditions and participants.

A network-level description is particularly suited to MS. Damage in MS is diffuse, involving white matter tracts and grey matter throughout the brain, and cognitive impairment is increasingly understood as a disturbance of brain networks rather than of single regions (Schoonheim et al., 2015, 2022). MEG studies in source space, mostly of resting-state activity, have shown altered functional networks in patients. Projecting MEG activity onto atlas regions with a beamformer, Tewarie et al. (2014a) found lower global integration of alpha2- and beta-band networks in early MS, related to poorer cognitive performance, and in a larger sample with long-standing MS functional network changes were related to the topology of grey matter networks (Tewarie et al., 2014b). A reduced individual specificity of source-reconstructed functional connectomes predicted fatigue (Cipriano et al., 2023), and a systematic review of MEG studies in MS reported reduced power modulations and connectivity (Khan et al., 2021). Evoked responses such as the P300 also arise from distributed generators (Polich, 2007), so that alterations that are weak at any single location may become detectable when activity is combined across the sources of a network.

Here we applied BROAD-NESS to MEG recordings of a three-stimulus auditory oddball in MS patients and healthy controls and compared the network-level result with a conventional source-by-source analysis of the same contrast. We asked whether the networks supporting auditory deviance and target processing differ between groups, with particular interest in the differentiation of task-relevant targets from task-irrelevant deviants. We also asked whether network measures relate to task performance, cognition and fatigue in patients.

## 2. Materials and methods

### 2.1 Participants and clinical assessment

Patients with multiple sclerosis (MS) and healthy controls (HC) were enrolled at the Centre for Diagnosis and Treatment of Demyelinating Diseases, University Hospital of Bari, Italy. The cohort overlaps with that of Paolicelli et al. (2021), who analysed sensor-level MEG and high-density EEG responses to a two-stimulus auditory oddball (240 standard and 60 target tones). The present study analyses a different task recorded in the same sessions, a three-stimulus auditory oddball. According to the acquisition dates stored in the recording files, the included participants were recorded between February 2018 and February 2020 (patients: March 2018 to July 2019), a period extending beyond the enrolment period reported by Paolicelli et al. (2021) (March 2018 to April 2019).

Twenty-two HC and 25 MS patients were enrolled, and three-stimulus oddball recordings were available for analysis for 21 HC and 23 MS patients. Participants were excluded only for problems of the recording file or of its preprocessing, never on the basis of the neural signal: stimulus trigger codes were absent from the recording (three HC, one MS patient); the recording file could not be read (two HC); the file stored for one HC was a copy of another participant’s recording and the original could not be retrieved; and Maxwell filtering returned no usable data for two MS patients (all data segments were rejected from the start of the recording in one case and after 88 s in the other; Section 2.3). No participant was excluded by the data-quality criteria described in Section 2.3. The final sample comprised 15 HC and 20 MS patients (participant flow in Supplementary Fig. S1).

The study protocol (Paolicelli et al., 2021) required age between 18 and 55 years and, for patients, a recent diagnosis of clinically isolated syndrome or relapsing-remitting MS according to the 2017 McDonald criteria (Thompson et al., 2018), a non-active phenotype (Lublin et al., 2014), minimal disability and no previous disease-modifying treatment. Exclusion criteria were disease duration above 5 years, Expanded Disability Status Scale (EDSS; Kurtzke, 1983) above 6, relapse or steroid treatment in the previous 30 days, psychiatric or other neurological disease, antidepressant or antipsychotic treatment, substance abuse, major head injury, moderate-to-severe hearing loss, and vascular or metabolic conditions. The clinical records of the included patients show deviations from some of these criteria (age above 55 years in two patients, disease duration above 5 years in three, previous disease-modifying treatment in two, antidepressant treatment in one and congenital hearing loss in one). All patients were included in the analyses, and the characteristics of the sample are reported in Table 1.

**Table 1.** Demographic and clinical characteristics of the sample. Values are mean ± SD, or median (range) for disease duration, EDSS and lesion count; n is given when data were incomplete. Disease duration is measured from symptom onset. FSS and PASAT-3 scores of zero were treated as missing.

| Characteristic | HC (n = 15) | MS patients (n = 20) |
|---|---|---|
| Age (years) | 36.4 ± 10.3 | 39.4 ± 10.7 (n = 16) |
| Sex (F/M) | 9/6 | 13/3 (n = 16) |
| Education (years) | 13.6 ± 3.1 (n = 13) | 12.9 ± 2.9 (n = 16) |
| Phenotype (CIS/RRMS/other) | – | 1/14/1 (n = 16) |
| Disease duration (years) | – | 1.8 (0.3–26.7) (n = 16) |
| EDSS | – | 2.0 (1.5–4.5) (n = 16) |
| FSS | 2.9 ± 1.5 (n = 6) | 2.5 ± 1.3 (n = 14) |
| BDI-II | 1.8 ± 3.5 (n = 13) | 5.0 ± 4.5 (n = 16) |
| SDMT | 53.2 ± 13.7 (n = 13) | 47.2 ± 13.9 (n = 16) |
| PASAT-3 | 42.6 ± 8.8 (n = 12) | 31.2 ± 12.9 (n = 13) |
| Cognitive impairment (≥ 2 altered tests), n | 2 of 13 | 4 of 16 |
| Total lesion count | – | 7.5 (2–16) (n = 16) |

All participants underwent a 45-min neuropsychological evaluation with Rao’s Brief Repeatable Battery of Neuropsychological tests (BRB-N), covering verbal memory, visuospatial memory, information processing speed and attention (Symbol Digit Modalities Test, SDMT; Paced Auditory Serial Addition Test, PASAT-3 and PASAT-5) and verbal fluency, together with the Trail Making Test. A test was considered altered when its z-score fell more than 2 standard deviations below the Italian normative mean (Amato et al., 2006), and cognitive impairment was defined as two or more altered tests (Paolicelli et al., 2021). Fatigue was assessed with the Fatigue Severity Scale (FSS; Krupp et al., 1989; cut-off 4.5) and depressive symptoms with the Beck Depression Inventory-II (BDI; Beck et al., 1996; cut-off 9). The total lesion count was the sum of juxtacortical/cortical, periventricular, infratentorial, spinal cord and optic nerve lesions on the baseline clinical MRI (median 3 months before the visit, range 1 day to 20 months). Clinical data were available for 16 of the 20 patients included. In HC, demographic data were available for all 15 participants, neuropsychological and BDI scores for 13 and FSS scores for 6. Demographic and clinical variables were compared between groups with Mann–Whitney U tests and, for sex and cognitive impairment, with Fisher’s exact test.

The study was conducted in accordance with the Declaration of Helsinki and approved by the Ethics Committee of the Azienda Ospedaliero-Universitaria Consorziale Policlinico di Bari (protocol code 5692, 20 June 2019). All participants gave written informed consent.

### 2.2 Task and MEG recording

Participants performed a three-stimulus auditory oddball task. Each session comprised 380 tones of 100 ms: 304 standard tones (250 Hz; 80%), 38 target tones (1000 Hz; 10%) and 38 non-target tones (625 Hz; 10%). Tones were presented binaurally through headphones at 75 dB, in pseudorandom order, with a stimulus onset asynchrony of 2–3 s; a session lasted about 16 min. Participants sat with eyes open and were instructed to press a button with the right hand as quickly as possible after each target tone and to ignore the other tones. Stimulus onsets were marked on the trigger channel. A second trigger, present for every stimulus, started 12–13 ms after the stimulus code. Latencies are reported relative to the stimulus code.

MEG was recorded with a 306-channel Elekta Neuromag TRIUX system (Elekta Neuromag Oy, Helsinki, Finland; 102 magnetometers and 204 planar gradiometers) in a dimly lit magnetically shielded room (ETS-Lindgren Euroshield, Eura, Finland), with active shielding (MaxShield) on. Data were sampled at 1000 Hz, the rate stored in the recording files (Paolicelli et al., 2021, report 1024 Hz), with an online band-pass filter of 0.1–330 Hz. Head position was tracked continuously with five head-position indicator coils, three on the forehead near the hairline and one over each mastoid.

Coil positions, the three anatomical fiducials, the EEG electrode positions and 96–152 additional head-shape points (median 107) were digitized before the recording. Vertical electro-oculogram (EOG) was recorded in all participants except one HC, and the electrocardiogram (ECG) in the other HC but not in patients. A 64-channel EEG was recorded simultaneously but is not analysed here.

### 2.3 Preprocessing, source reconstruction and epoch rejection

Raw data were processed with MaxFilter 2.2.15 (Elekta Neuromag) using temporal signal space separation (tSSS; Taulu and Simola, 2006) with a 4-s buffer and a subspace correlation limit of 0.98, fine-calibration and cross-talk correction, an SSS expansion origin at (0, 0, 40) mm in head coordinates, internal and external expansion orders of 8 and 3 (80 and 15 components), and movement compensation based on continuous head-position estimates from the five coils. Automatic bad-channel detection was off. Data were downsampled by a factor of 4 to 250 Hz, with an anti-aliasing low-pass at 83.3 Hz applied by MaxFilter.

Data were then converted to SPM format and processed in MATLAB (MathWorks, Natick, MA, USA) with custom code (LBPD, https://github.com/leonardob92/LBPD-1.0) and the OHBA Software Library (OSL; Woolrich et al., 2011), which integrates functions from FieldTrip (Oostenveld et al., 2011), FSL (Woolrich et al., 2009) and SPM (Penny et al., 2011). Independent component analysis (ICA; FastICA, Hyvärinen, 1999) was applied to the continuous data, and components reflecting eye blinks, eye movements and heartbeat were identified and removed. Sixty-two components were estimated per participant; components correlating with the EOG or ECG channel (|r| > 0.3) were flagged automatically, and all components were then inspected for topography and time course before removal (in patients, without ECG, cardiac components were identified from topography and time course only). The number of removed components, obtained from the reduction in data rank, was 2.6 ± 0.9 (range 2–5) in HC and 3.4 ± 1.5 (range 2–7) in MS patients (Welch t-test, p = 0.08). For the HC without EOG and ECG recordings, artefactual components were identified from topography and time course alone; this participant was included in the sensitivity analysis (Section 2.5).

Trigger values were read as unsigned 16-bit integers, and stimulus codes were taken from the low-order bits, retaining one event per stimulus. This step was needed because the response-pad bit was stored as a negative value in some recordings and because the second trigger could overlap with the stimulus code. Epochs from −100 to 3400 ms around stimulus onset were extracted and baseline-corrected (−100 to 0 ms). The number of extracted epochs was 380 in 20 participants, 378 or 379 in 11 and between 239 and 370 in four (median 380).

Individual structural MRIs were not available, so the same MNI152 template (2-mm T1) served as head model for all participants. The template scalp surface was extracted with FSL (FLIRT, BET and BETSURF; Jenkinson et al., 2012), and each participant’s digitized fiducials and head-shape points were coregistered to it with the RHINO procedure of OSL: initial alignment on the three fiducials, then refinement by iterative closest point on the head-shape points with 10 random starts. A single-shell forward model (Nolte, 2003) was computed for 3559 sources on an 8-mm grid within the MNI152 brain mask.

Source time series were estimated with a linearly constrained minimum variance (LCMV) beamformer (Van Veen et al., 1997) using the 102 magnetometers. Each sensor’s time series was z-scored within each epoch, and the data covariance matrix was computed over all epochs of the three conditions and over the full epoch (−100 to 3400 ms). The covariance was inverted with a pseudo-inverse restricted to its first 50 principal components, to account for the reduced rank of tSSS-processed data. For each source, the lead field was reduced to the orientation of maximum output power (singular value decomposition), and the weights were normalized by their norm (W/√(WWᵀ)) to reduce the bias towards the centre of the head (Huang et al., 2004). A single set of weights per participant was applied to the condition-averaged evoked fields, and source time series were baseline-corrected (−100 to −4 ms).

Epochs contaminated by artefacts not removed by ICA, such as movement related to the button press, were excluded from the evoked averages. Detection used the ICA-cleaned magnetometer data in the analysis window (−100 to 900 ms), low-pass filtered at 40 Hz and linearly detrended for this purpose only. An epoch was rejected if the peak-to-peak amplitude exceeded 4000 fT on any magnetometer, or if all magnetometers were flat (< 1 fT). Condition averages were recomputed from the retained epochs and projected through the same beamformer weights. Recomputing the averages from all epochs reproduced the stored source time series exactly in every participant (r = 1.0), confirming that this step used the original beamformer.

Two exclusion criteria were set before the group data were examined: fewer than 20 retained target or non-target epochs, and baseline noise (root mean square across magnetometers, −100 to 0 ms, median over retained epochs) above the sample median + 3 scaled median absolute deviations. Criteria were based on data quality only. The signal-to-noise ratio of evoked responses was not used for exclusion, because it depends on response amplitude, which may itself be altered by the disease.

### 2.4 Broadband network estimation

All analyses used the interval from −100 to 900 ms (251 samples), which excludes the response to the following stimulus. Because the sign of beamformer time series is arbitrary for each source and participant, signs were aligned across participants before averaging. For each source and participant, the sign was inverted if the covariance between that participant’s time course (0–600 ms, three conditions concatenated, mean removed) and the average of all other participants was negative. The procedure was iterated until fewer than 0.5% of source-by-participant pairs changed sign. It did not use group labels, and the same sign was applied to all conditions, so that condition contrasts were unaffected.

Brain networks were estimated with the BROAD-NESS (Bonetti et al., 2025a, 2025b; Andersen et al., 2026; Christensen et al., 2026; Fernández-Rubio et al., 2026) analytical pipeline developed in the larger NESS project (Rosso et al., 2025; Shinozuka et al., 2025, 2026; Kudriavtsev et al., 2026; Malvaso et al., 2026). Source time series were averaged over all participants and then over the three conditions, and principal component analysis (PCA) was applied to the resulting sources × time matrix, with time points as observations. Each component’s eigenvector defines the spatial activation pattern of a network (sign set so that the largest absolute weight is positive). Network time series for each participant and condition were obtained by projecting the participant’s source time series onto each eigenvector. Networks were retained according to the elbow of the explained-variance curve; effective dimensionality was computed as the participation ratio of the eigenvalue spectrum. For anatomical description, the 5% of sources with the largest absolute weight in each network were labelled with the Automated Anatomical Labeling atlas (Tzourio-Mazoyer et al., 2002).

### 2.5 Statistical analysis

Analysis windows were defined for each network with a collapsed localizer (Luck and Gaspelin, 2017), computed on the average of the two group means so that both groups contributed equally. Two localizer signals were used: the mean across the three conditions, searched between 30 and 300 ms, and the deviant-minus-standard difference ((target + non-target)/2 − standard), searched between 150 and 700 ms. After smoothing with a 20-ms moving average, local extrema exceeding 25% of the maximum absolute amplitude and at least 40 ms apart were selected. Windows were centred on each extremum (±20 ms for peaks before 150 ms, ±50 ms thereafter), and overlapping windows were merged. Windows are named by network and latency. The localizer is independent of the group contrast and also of the target-versus-non-target contrast, whose condition weights are orthogonal to those of the deviant-minus-standard signal.

For each participant, network and window, mean amplitudes were computed for each condition and for three planned contrasts: target − standard, non-target − standard and target − non-target. The last contrast compares two equally rare deviants that differ in task relevance and addresses the main question of the study (Section 1); the other two characterize the response to deviance. Groups were compared with Welch’s t-test; effect sizes are reported as Hedges’ g with 95% confidence intervals (Hedges and Olkin, 1985), and evidence for or against a group difference as a Jeffreys–Zellner–Siow Bayes factor (BF10; Cauchy prior scale √2/2; Rouder et al., 2009). Condition effects were tested across all participants with one-sample t-tests on the contrasts. Window-based tests are reported without correction for multiple comparisons and interpreted together with effect sizes and Bayes factors; inference on group differences rests on the cluster-based tests below.

For each network and planned contrast, groups were compared at each time point between 0 and 900 ms with Welch’s t-statistic. Adjacent time points with |t| above the two-tailed critical value (t(33) = 2.03) and the same sign formed clusters, whose mass was the sum of their t-values (Maris and Oostenveld, 2007). This cluster-forming threshold corresponds to an uncorrected two-tailed p <0.05 at each time point; because the degrees of freedom of Welch’s test vary with the data, a single threshold based on the pooled degrees of freedom was used for all time points and permutations. The threshold affects the sensitivity of the test to effects of different extent but not its control of the family-wise error rate (Maris and Oostenveld, 2007). The null distribution was obtained from 5000 random permutations of group labels (fixed random seed), taking in each permutation the maximum absolute cluster mass across the four networks. This controls the family-wise error rate across networks within each contrast (p = (k + 1)/5001, where k is the number of permutations with a mass at least as large as the observed one). Because three contrasts were tested, a more conservative correction across all contrasts and networks, using the maximum over the twelve tests in each permutation, is also reported.

Group differences in latency were tested with the jackknife procedure (Miller et al., 1998; Ulrich and Miller, 2001; Kiesel et al., 2008). Within each window, latency was the time at which 50% of the area under the response, rectified with the polarity of the localizer peak, was reached (linear interpolation between samples). Jackknife standard errors were combined across groups into a t-statistic with 33 degrees of freedom. For windows defined only on the deviant-minus-standard signal, latencies were computed for target and non-target responses only.

The main result was examined in five ways: (i) leave-one-participant-out estimation of the effect; (ii) re-estimation of the networks from the average of the two group means, so that groups were weighted equally; (iii) comparison between groups of baseline noise in each network and condition, of the signal-to-noise ratio of the N1m, of the number of retained epochs and of the number of removed ICA components, which was also entered as a covariate of the group effect; (iv) a sensitivity analysis excluding four participants from all statistics (networks were still estimated on the full sample). They were chosen because each could directly affect the measured auditory responses or their cleaning: a patient taking central nervous system-active medication (an antidepressant and an antiepileptic), a patient with congenital hearing loss, a patient with exactly 20 retained target epochs, and the HC without EOG and ECG recordings. Other deviations from the protocol criteria (age, disease duration, previous treatment) were not grounds for exclusion. (v) A second sensitivity analysis, added after inspection of the behavioural data, excluded the three participants who pressed the button after most non-target tones (Section 3.1), for whom non-targets may also have been task-relevant.

To compare the network-level result with a conventional source-level analysis and to describe where in source space the main effect arose, the target − non-target amplitude in the localizer window of network 2 (378–626 ms) was computed for every source after sign alignment and compared between groups with Welch’s t-test. Each source was oriented by the direction of the mean response of the two groups, so that positive values indicate a smaller difference in patients. Clusters of adjacent sources (26-neighbourhood on the 8-mm grid) with |t| > 2.03 were tested with 5000 permutations of group labels, controlling the family-wise error rate across all sources.

### 2.6 Behaviour and relationships with clinical measures

Button presses were read from the trigger channel of the Maxwell-filtered recordings (1000 Hz). A press was assigned to a stimulus if it occurred 150–1500 ms after stimulus onset. Hits, misses, false alarms to standards and false alarms to non-targets were counted, and median reaction times were computed for hits. Discrimination between targets and non-targets was quantified as d′ with a log-linear correction (Hautus, 1995). Button presses were not recorded for one HC and three MS patients. Groups were compared with Mann–Whitney U tests.

Within MS patients, two relationships were specified before the analysis, using Spearman’s correlations and localizer windows rather than cluster windows to avoid circular selection: (H1) target − non-target amplitude in network 2 (378–626 ms) with d′ and with the false-alarm rate to non-targets; (H2) the P3 response of network 1 (target − standard, 310–742 ms) with FSS, as a source-level test of the association reported by Paolicelli et al. (2021). Correlations of the same two measures with hit rate, reaction time, EDSS, disease duration, BDI, SDMT, PASAT-3, number of altered cognitive tests and lesion count were exploratory and corrected with the Benjamini–Hochberg false discovery rate (Benjamini and Hochberg, 1995). FSS and PASAT-3 values of zero were treated as missing.

Analyses were run with MaxFilter 2.2.15 and MATLAB R2022a, using LBPD, OSL (Woolrich et al., 2011), FieldTrip (Oostenveld et al., 2011), SPM (Penny et al., 2011), the BROAD-NESS toolbox (Bonetti et al., 2025a) and custom scripts (see Data and code availability).

## 3. Results

### 3.1 Sample, data quality and behaviour

The final sample comprised 15 HC and 20 MS patients (Table 1). The groups did not differ in age, sex, education, fatigue, SDMT score or proportion of participants with cognitive impairment (all p ≥ 0.25), whereas MS patients had higher BDI scores (p = 0.008) and lower PASAT-3 scores (p = 0.022) (Table 1; Supplementary Table S8). After epoch rejection, HC retained on average 293.1 standard (range 190–304), 36.8 target (23–38) and 37.1 non-target epochs (26–38), and MS patients 298.4 (260–304), 36.2 (20–38) and 36.7 (29–38). The number of retained target and non-target epochs did not differ between groups (p = 0.67 and p = 0.72). No participant met the exclusion criteria for data quality. ICA removed 2.6 ± 0.9 components in HC and 3.4 ± 1.5 in MS patients (p = 0.08). Sign alignment inverted 33.1 ± 4.2% of sources in HC and 31.9 ± 4.5% in MS patients (Welch t = −0.82, p = 0.42). Baseline noise did not differ between groups in any network or condition (all p ≥ 0.19), nor did the signal-to-noise ratio of the N1m (median 31.0 in HC and 25.6 in MS patients; p = 0.97) (Supplementary Table S5).

Task performance was recorded in 14 HC and 17 MS patients and did not differ between groups (Table 2). Hit rates were high in both groups (median 0.97), and median reaction times were 565 ms in HC and 558 ms in MS patients (p = 0.38). Three participants (one HC and two MS patients) detected 37 of 38 targets but also pressed the button after 33 to 38 of the 38 non-target tones, which lowered their d′; excluding them did not change the main result (Section 3.4).

**Table 2.** Behavioural performance (median [interquartile range]; Mann–Whitney U test).

| Measure | HC (n = 14) | MS patients (n = 17) | p |
|---|---|---|---|
| Hit rate | 0.97 [0.95–1.00] | 0.97 [0.91–1.00] | 0.93 |
| Reaction time (ms) | 565 [399–614] | 558 [498–763] | 0.38 |
| False alarms to non-targets (rate) | 0.04 [0.00–0.13] | 0.08 [0.05–0.22] | 0.12 |
| d′ (target vs non-target) | 3.41 [2.84–4.00] | 2.72 [2.34–3.57] | 0.20 |

### 3.2 Broadband networks

The first four components explained 48.8%, 31.3%, 10.6% and 3.3% of the variance (94.0% in total), after which the explained-variance curve flattened; effective dimensionality was 2.86 (Fig. 2). Anatomical labels refer to the 5% of sources with the largest absolute weight (Supplementary Table S1); with an 8-mm grid and a template head model they indicate approximate locations. Network 2 loaded on bilateral perisylvian cortex (postcentral gyrus, Rolandic operculum, insula, superior temporal gyrus and left supramarginal gyrus), with opposite signs in the two hemispheres. Network 4 covered a similar bilateral perisylvian and sensorimotor territory with the same sign in both hemispheres (postcentral and left precentral gyrus, Rolandic operculum, insula), together with the left thalamus. Network 1 was right-lateralized, with its largest weights in the right hippocampus and parahippocampal gyrus, mid-cingulate cortex, supplementary motor area, precentral and superior temporal gyri. Network 3 loaded mainly on right deep and temporal structures (caudate, putamen, thalamus, parahippocampal and inferior temporal gyri), and 40 of its strongest sources fell outside the AAL labels. Labels of deep and medial temporal sources should be read with caution, given the limited sensitivity of MEG to deep activity. Network 2 showed the auditory evoked sequence, with a large N1m peaking at 100–106 ms for all three tones and larger for the two deviants, followed by a sustained response to the deviants between about 160 and 630 ms (Fig. 3). Network 1 showed early responses at 80–91 ms and a large sustained deflection that began at about 250 ms, peaked at about 350–400 ms and lasted until about 800 ms; it was present for target and non-target tones, larger for targets, and small for standards, with the time course of the P3 complex. Network 3 responded at about 45 and 90 ms and showed a deviant-specific deflection between about 190 and 300 ms, larger for targets. Network 4 responded to all tones at about 75 and 130 ms; between about 170 and 260 ms the response to standards had the opposite polarity to that to deviants, and after about 300 ms target and non-target responses had opposite polarities.

**Figure 1.**
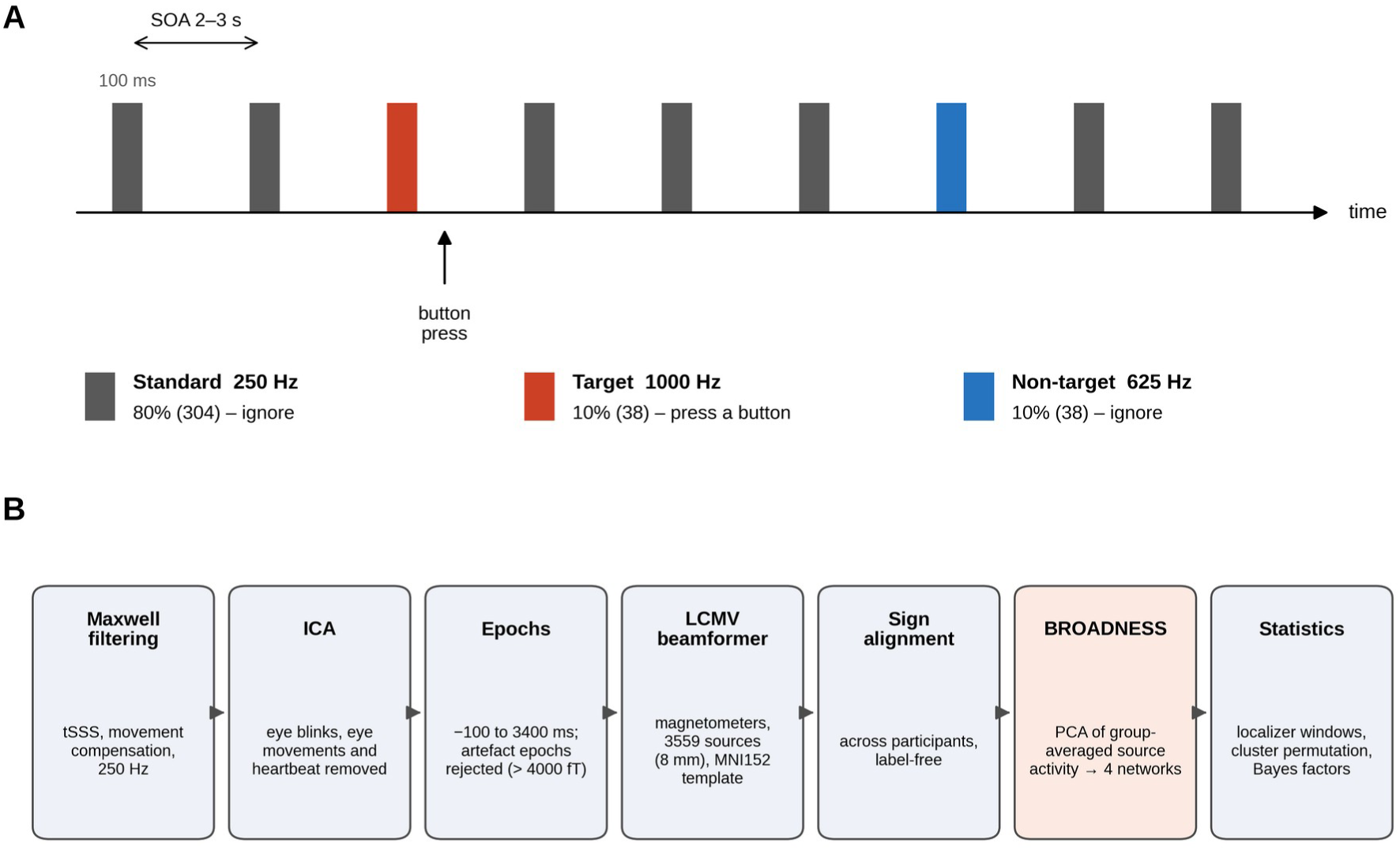
Experimental paradigm and analysis pipeline. (A) Three-stimulus auditory oddball: standard tones (250 Hz, 80%), targets (1000 Hz, 10%) requiring a button press, and non-targets (625 Hz, 10%) to be ignored; tone duration 100 ms, stimulus onset asynchrony 2–3 s. (B) Processing steps: Maxwell filtering with movement compensation, ICA, epoch rejection, LCMV beamforming on an 8-mm grid, sign alignment across participants, network estimation with BROAD-NESS, and statistical analysis.

**Figure 2.**
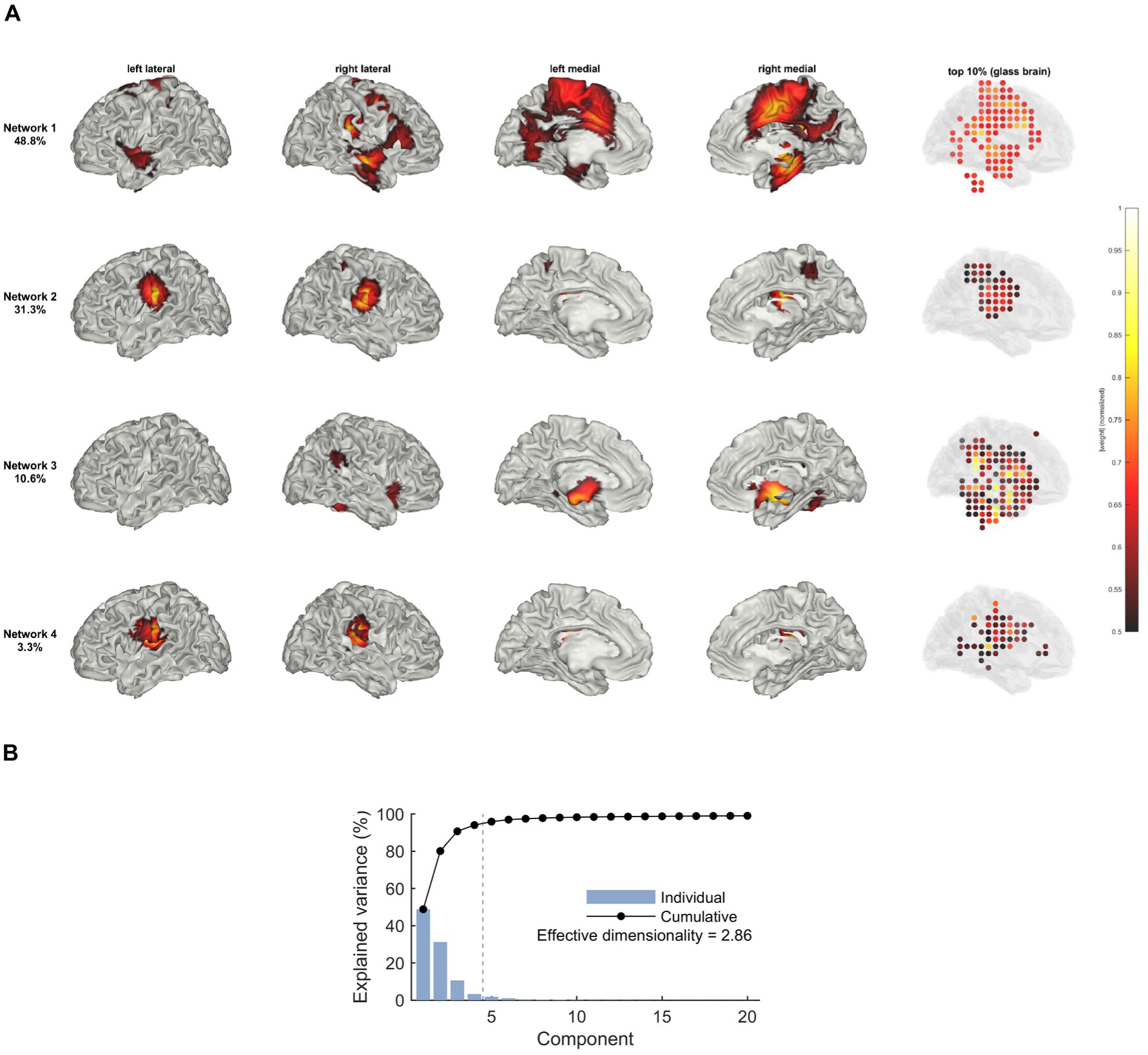
Broadband brain networks. (A) Absolute spatial weights of the four retained networks, normalized to the maximum of each network and projected on the MNI cortical surface (left and right lateral and medial views; each vertex shows the Gaussian-weighted mean (σ = 6 mm) of sources within 16 mm; weights above 50% of the maximum). The glass brain on the right shows the sources that are among the 10% with the largest weights and exceed 50% of the maximum, including deep sources that the surface cannot show. Percentages give the variance explained by each network. The projection interpolates data on an 8-mm grid, whose effective resolution is limited by MEG spatial resolution (1–2 cm). (B) Variance explained by the first 20 components and cumulative variance; the dashed line marks the four retained networks. Signed maps are shown in Supplementary Fig. S3.

**Figure 3.**
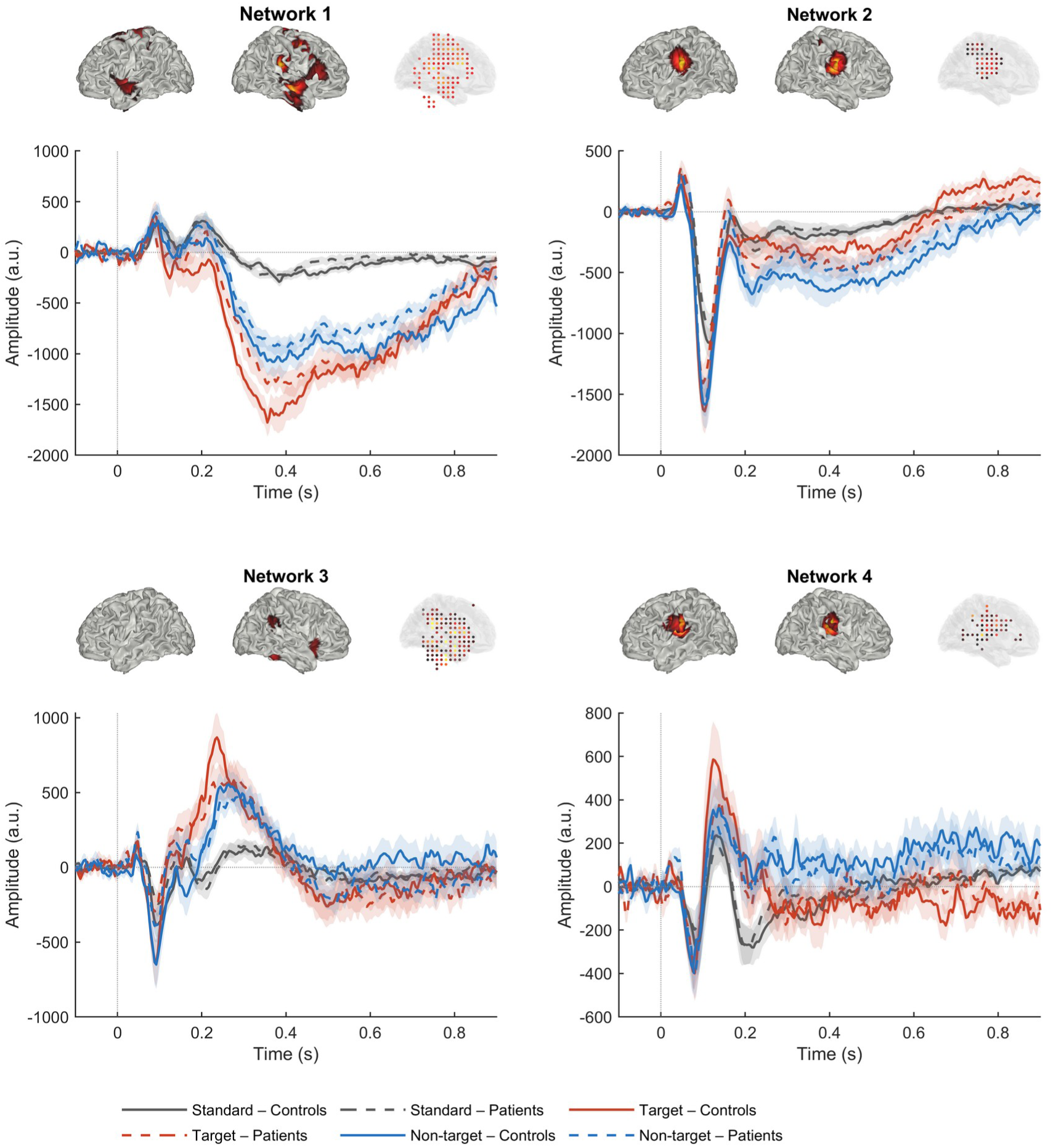
Time series of the four networks for standard (grey), target (red) and non-target (blue) tones in HC (solid lines) and MS patients (dashed lines). Shaded areas: standard error of the mean. Amplitudes are in arbitrary units; the sign of each network is arbitrary. Above each panel, the spatial pattern of the network (left and right lateral views and glass brain; same threshold and colour scale as in Fig. 2A).

### 3.3 Condition effects and window-based group comparisons

Eleven analysis windows were identified across the four networks (Supplementary Table S2). Deviant tones differed from standards in all four networks: the target − standard and non-target – standard contrasts were significant across participants in every window after 110 ms (all p ≤ 0.03), except target − standard in network 4 between 294 and 394 ms (p = 0.62), and target and non-target responses differed in network 1 (310–742 ms), network 2 (162–370 and 378–626 ms), network 3 (186–298 ms) and network 4 (294–394 ms) (all p ≤ 0.002) (Supplementary Table S3).

Of the 66 group comparisons computed in the analysis windows, four had uncorrected p < 0.05. Two concerned the target − non-target contrast in network 2: between 162 and 370 ms (Hedges’ g = −0.78 [−1.47, −0.08], p = 0.034, BF10 = 2.5) and between 378 and 626 ms (g = −1.11 [−1.82, −0.39], p = 0.003, BF10 = 16.1). In both windows HC responded differently to target and non-target tones (mean difference 258 and 297 a.u.), whereas MS patients did not (94 and 41 a.u.) (Fig. 4E). The other two concerned the non-target response of network 3 between 28 and 68 ms (non-target: g = 0.68, p = 0.046; non-target − standard: g = 0.76, p = 0.027, BF10 = 2.3), a window preceding the N1m in which condition effects were absent (p ≥ 0.46). N1m and P3 amplitudes did not differ between groups in any condition (all p ≥ 0.13; BF01 between 1.1 and 2.9), and no group difference in latency was found in any window or condition (all jackknife p ≥ 0.13; Supplementary Table S4).

**Figure 4.**
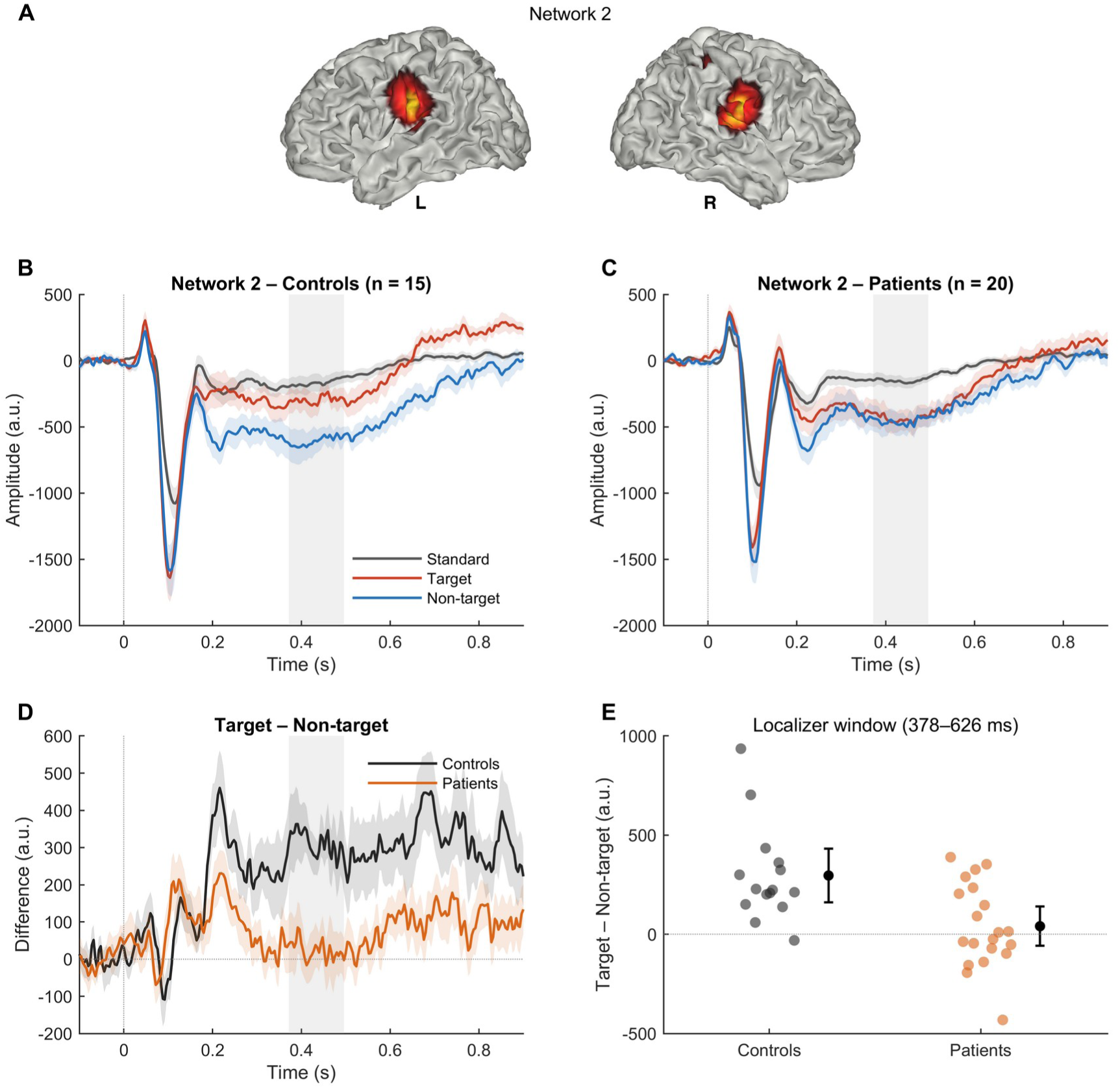
Reduced differentiation of target and non-target tones in network 2 in MS patients. (A) Spatial pattern of network 2 (left and right lateral views; as in Fig. 2A). (B, C) Network 2 time series by condition in HC (n = 15) and MS patients (n = 20); same scale in both panels. (D) Target − non-target difference by group (mean ± SEM). The grey band marks the cluster with a significant group difference (372–496 ms; p = 0.020, family-wise corrected across the four networks; p = 0.108 when correcting across all three contrasts and four networks). (E) Individual target − non-target amplitudes in the localizer window (378–626 ms), with group means and 95% confidence intervals; Hedges’ g = −1.11 [−1.82, −0.39], uncorrected p = 0.003, BF10 = 16.1.

### 3.4 Cluster-based group comparisons

The cluster-based tests identified one group difference: in network 2, the target − non-target contrast differed between 372 and 496 ms (cluster mass −96.5; p = 0.010 for network 2, p = 0.020 corrected across the four networks) (Fig. 4D). In this interval the mean difference between target and non-target responses was 313 a.u. in HC and 24 a.u. in MS patients. No other cluster reached significance for any contrast or network (all corrected p ≥ 0.24; Table 3). When the correction extended to all three contrasts and four networks, the p-value of this cluster was 0.108.

**Table 3.** Cluster-based group comparisons (MS patients vs HC, 0–900 ms). For each contrast and network, the cluster with the largest absolute mass is shown. Corrected p controls the family-wise error across the four networks within each contrast; across all three contrasts and four networks, the network 2 target − non-target cluster had p = 0.108.

| Contrast | Network | Cluster (ms) | Mass | p (network) | p (corrected) |
|---|---|---|---|---|---|
| Target − standard | 1 | 328–344 | 10.8 | 0.361 | 0.841 |
| Target − standard | 2 | 8–20 | 12.7 | 0.365 | 0.778 |
| Target − standard | 3 | 856 | −2.1 | 0.789 | 0.996 |
| Target − standard | 4 | 16–20 | −4.5 | 0.693 | 0.973 |
| Non-target − standard | 1 | 776–796 | 20.5 | 0.158 | 0.498 |
| Non-target − standard | 2 | 60–72 | 11.3 | 0.360 | 0.808 |
| Non-target − standard | 3 | 616–640 | −16.0 | 0.233 | 0.636 |
| Non-target − standard | 4 | 28–48 | 14.8 | 0.245 | 0.677 |
| Target − non-target | 1 | 764–796 | −24.2 | 0.102 | 0.365 |
| Target − non-target | 2 | 372–496 | −96.5 | 0.010 | **0.020** |
| Target − non-target | 3 | 112 | −2.1 | 0.872 | 1.000 |
| Target − non-target | 4 | 16–36 | −13.7 | 0.279 | 0.728 |

The effect was robust (Supplementary Table S6). Removing one participant at a time left the effect size between g = −1.23 and −1.02 (p between 0.002 and 0.006). Networks estimated with equal weight for the two groups were nearly identical to the original ones (|r| ≥ 0.99 for all four networks) and gave the same result (g = −1.10, p = 0.003; cluster 372–496 ms, p = 0.006 for network 2). Excluding the four participants of the sensitivity analysis (14 HC, 17 MS patients) left the cluster at 376–464 ms (p = 0.041 corrected across networks) and the effect in the localizer window unchanged (g = −1.16 [−1.92, −0.40], p = 0.003, BF10 = 14.4) (Supplementary Fig. S5). Excluding the three participants who also responded to non-target tones (14 HC, 18 MS patients) did not change the result (cluster 368–500 ms, p = 0.016 corrected across networks; localizer window g = −1.22 [−1.97, −0.46], p = 0.002, BF10 = 22.5) (Supplementary Table S6). The number of removed ICA components was unrelated to the effect (Spearman ρ = −0.15, p = 0.39), and the group difference remained when it was entered as a covariate (−233 a.u., t = −2.89, p = 0.007).

At the level of single sources, no spatial cluster survived correction (largest cluster: p = 0.35). The largest uncorrected clusters both indicated a smaller difference in patients and lay in left postcentral, precentral, inferior parietal and middle temporal regions and in right postcentral, mid-cingulate and inferior parietal regions, overlapping the sensorimotor and perisylvian part of network 2 (Supplementary Fig. S7, Table S9). The group difference was therefore expressed at the level of the network rather than in a circumscribed region.

### 3.5 Relationships with behaviour and clinical measures

In MS patients, the target − non-target amplitude of network 2 was not related to discrimination between targets and non-targets (d′: ρ = −0.04, p = 0.88; false-alarm rate to non-targets: ρ = −0.09, p = 0.75; n = 17), and the P3 amplitude of network 1 was not related to fatigue (FSS: ρ = −0.20, p = 0.49; n = 14). Among the exploratory correlations, none survived correction for multiple comparisons (all q ≥ 0.32). The largest were between the network 2 target − non-target amplitude and the SDMT (ρ = −0.54, uncorrected p = 0.030) and the number of altered cognitive tests (ρ = 0.54, p = 0.032; n = 16) (Supplementary Table S7).

## 4. Discussion

We applied BROAD-NESS to MEG recordings of a three-stimulus auditory oddball in MS patients and healthy controls. Four broadband networks captured the main spatiotemporal dynamics of the response in both groups and were strongly modulated by stimulus type. Early auditory responses and the P3 complex did not differ between groups. The one group difference concerned network 2, which involved bilateral perisylvian and sensorimotor regions: between 370 and 500 ms, controls responded differently to target and non-target tones, whereas patients did not. This difference was observed although task performance was comparable in the two groups.

### 4.1 Reduced differentiation of task-relevant and task-irrelevant deviants

In controls, the late sustained response of network 2 was larger for non-targets than for targets, so that two equally rare deviants were processed differently according to their task relevance. In MS patients the two responses were similar, and neither condition alone differed significantly between groups: the difference lay in their differentiation. The effect was large (g = −1.11 in the localizer window), was supported by a Bayes factor of 16 and survived correction across networks in the cluster test. It was stable when single participants were removed, when the networks were estimated with equal weight for the two groups and when the number of removed ICA components was taken into account, and it remained when the participants with particular clinical or recording characteristics, or those who also responded to non-target tones, were excluded.

The effect emerged after the N1m, during the P3 interval, while early auditory responses were preserved. This timing suggests that the difference arises at stages of attentional selection and stimulus evaluation rather than of initial sensory encoding, and it fits a stage at which attention is redirected after a distractor (Schröger and Wolff, 1998). A reduced differentiation could therefore reflect weaker modulation of auditory processing by task relevance, in line with the deficits of attention and processing speed that characterize cognitive impairment in MS (Chiaravalloti and DeLuca, 2008). Target and non-target tones, however, also differed in frequency and in their acoustic distance from the standard (1000 vs 625 Hz, standard 250 Hz), so that a reduced sensitivity to the magnitude of deviance is an alternative account. The present design cannot separate the two, although the absence of group differences in the earlier, acoustically driven responses makes a purely sensory explanation less likely. Designs that counterbalance the assignment of tones to the target and non-target roles would resolve this question.

### 4.2 Preserved sensory and P3 responses

N1m and P3 responses did not differ between groups in amplitude or latency, with Bayes factors mildly favouring the absence of a difference. This agrees with the similar P300 amplitude and topography found in an overlapping cohort with a two-stimulus task (Paolicelli et al., 2021) and extends it to the source level. The slight, non-significant latency prolongation reported there was not observed here; the tasks, the measures (scalp peaks vs network time series) and the latency methods differ, and Bayes factors close to 3 indicate that the data were only weakly informative about the absence of differences.

Network 1, whose late response had the time course of the P3 complex, showed a non-significant tendency towards a smaller non-target response in patients (310–742 ms: g = 0.54, uncorrected p = 0.13), whereas its differentiation between target and non-target tones did not differ between groups (p = 0.68). Given the sample size, smaller alterations in this network cannot be excluded, and this observation is reported as descriptive only. Together, these results suggest that alterations of auditory processing in MS may not be expressed uniformly across networks but may preferentially affect specific aspects of late stimulus processing.

### 4.3 Neural differences in the absence of behavioural deficits

Patients detected targets as accurately and as fast as controls, and within patients the network 2 differentiation was not related to d′ or to false alarms to non-targets. The network difference was therefore not accompanied by a behavioural deficit, and the similar reaction times make it unlikely that it reflects differences in the timing of the motor response, which overlaps the late time window. Neural differences may thus be detectable while performance is preserved. Cross-sectional data cannot establish whether such differences precede cognitive decline or reflect compensatory processing, and the simple task, with high accuracy in both groups, may have been insensitive to subtle behavioural differences.

The network 1 P3 response was not related to fatigue. The inverse correlation between P300 amplitude and fatigue reported by Paolicelli et al. (2021), computed across patients and controls with a different task, was therefore not reproduced at the source level. The strongest exploratory correlations, with SDMT and with the number of altered tests, did not survive correction and pointed in an unexpected direction; they are not interpreted. Longitudinal studies and more demanding attentional tasks are needed to establish whether network measures of this kind have predictive or clinical value.

### 4.4 Contribution of network-level analysis

The main effect was identified at the network level, whereas a conventional mass-univariate analysis of the same contrast over the 3559 sources found no spatial cluster that survived correction (largest cluster: p = 0.35); the largest uncorrected clusters overlapped the sensorimotor and perisylvian sources of network 2. By combining sources that share a common time course, the network representation concentrates a distributed effect into a single time series and reduces the number of comparisons (Bonetti et al., 2025a). This observation does not show that network-level analysis is more sensitive than other source-analysis approaches in general, but it supports its use for subtle, distributed changes in clinical populations, in line with the view of MS as a disorder of brain networks (Schoonheim et al., 2022).

### 4.5 Limitations and future directions

The sample is small (15 HC and 20 MS patients), which limits power for smaller effects in other networks and for relationships with clinical and cognitive measures; effect sizes selected by statistical significance also tend to be overestimated. The main result survived correction across networks within the target − non-target contrast, but not the more conservative correction across all three contrasts (p = 0.108), and should be considered preliminary until it is replicated. Source reconstruction used a template head model, because individual MRIs were not available, which limits anatomical precision, particularly for deep sources; the beamformer used magnetometers only, and lesions were not modelled. Networks were estimated at the group level, so individual differences in their spatial layout are not captured.

Some patients did not meet all the inclusion criteria of the original protocol (age, disease duration, previous treatment, antidepressant use or hearing loss; Section 2.1). They were retained, so the sample is more heterogeneous than the protocol implies; excluding the patients whose medication or hearing could most directly affect auditory responses did not change the main result. Behavioural data were missing for four participants and clinical data for four patients. Finally, the acoustic differences between target and non-target tones and the button press required for targets are potential confounds that future designs should control.

### 4.6 Conclusions

In this sample of MS patients, the source-level networks engaged by an auditory three-stimulus oddball were largely preserved, including the sensory N1m and the P3 complex. A distributed auditory network, however, differentiated less between task-relevant and task-irrelevant deviants in the late interval, in the absence of differences in task performance. This selective alteration is a candidate marker of changes in attention-dependent auditory processing in MS, to be tested in larger and longitudinal samples.

## Data Availability

The code used for preprocessing and source reconstruction (LBPD, https://github.com/leonardob92/LBPD-1.0) and for network estimation (BROAD-NESS, https://github.com/leonardob92/BROADNESS_MEG_AuditoryRecognition) is publicly available. The study-specific analysis scripts are available from the corresponding authors on reasonable request. The MEG and clinical data cannot be shared publicly because they contain clinical information on patients; they are available from the corresponding authors on reasonable request.

https://github.com/leonardob92/BROADNESS_MEG_AuditoryRecognition

## CRediT authorship contribution statement

**Francesco Carlomagno:** Conceptualization, Data curation, Formal analysis, Methodology, Software, Validation, Visualization, Writing – original draft, Writing – review & editing. **Leonardo Bonetti:** Methodology, Software, Writing – original draft, Writing – review & editing. **Mattia Rosso:** Methodology, Software, Writing – original draft, Writing – review & editing. **Elvira Brattico:** Conceptualization, Funding acquisition, Methodology, Project administration, Supervision, Writing – original draft, Writing – review & editing. **Paolo Taurisano:** Conceptualization, Investigation, Writing – review & editing. **Madia Marika Biasi:** Investigation, Writing – review & editing. **Giusy Tancredi:** Investigation, Writing – review & editing. **Alessia Manni:** Data curation, Investigation, Writing – review & editing. **Antonio Iaffaldano:** Data curation, Investigation, Writing – review & editing. **Giulia Paparella:** Investigation, Writing – review & editing. **Daphne Gasparre:** Investigation, Writing – review & editing. **Marina de Tommaso:** Conceptualization, Funding acquisition, Methodology, Project administration, Resources, Supervision, Writing – review & editing. **Vitoantonio Bevilacqua:** Conceptualization, Funding acquisition, Methodology, Project administration, Resources, Supervision, Writing – review & editing. **Damiano Paolicelli:** Conceptualization, Funding acquisition, Methodology, Project administration, Resources, Supervision, Writing – review & editing.

## Acknowledgements

We thank all the patients and healthy volunteers who took part in the study.

## Declaration of generative AI and AI-assisted technologies in the manuscript preparation process

During the preparation of this work the authors used Claude (Anthropic) to assist with drafting and revising the text, including the description of materials and methods. After using this tool, the authors reviewed and edited the content as needed and take full responsibility for the content of the published article.

## Supplementary material (separate file)

| Item | Content |
|---|---|
| Fig. S1 | Participant flow: participants enrolled, recordings available, exclusions with reasons, final sample |
| Fig. S2 | Localizer signals and analysis windows for each network |
| Fig. S3 | Signed spatial patterns of the four networks |
| Fig. S4 | Target − non-target difference in all four networks, by group, with significant clusters |
| Fig. S5 | Sensitivity analysis: Fig. 4 without the four participants listed in Section 2.5 |
| Fig. S6 | A priori brain–behaviour and brain–fatigue relationships in MS patients |
| Fig. S7 | Source-level map of the group difference in target − non-target (378–626 ms), descriptive |
| Table S1 | Anatomical labels of the four networks (AAL, top 5% of sources) |
| Table S2 | Analysis windows per network and the localizer signal that defined them |
| Table S3 | All window-based statistics: condition effects and group comparisons |
| Table S4 | Latencies (jackknife) per network, window and condition |
| Table S5 | Data quality by group |
| Table S6 | Robustness of the main result |
| Table S7 | Correlations with behaviour and clinical measures |
| Table S8 | Neuropsychological scores by test in HC and MS patients |
| Table S9 | Source-level clusters |

