## Supplementary Materials for "Altered Auditory Network Differentiation in Multiple Sclerosis: A BROAD-NESS MEG Study"

### Supplementary Material

*Reduced differentiation between task-relevant and task-irrelevant deviant sounds in a source-level auditory network in multiple sclerosis: a MEG study*

HC: healthy controls (n = 15). MS: patients with multiple sclerosis (n = 20). a.u.: arbitrary units. Networks are numbered as in the main text; the sign of network weights and time series is arbitrary.

#### Supplementary figures

##### Figure S1

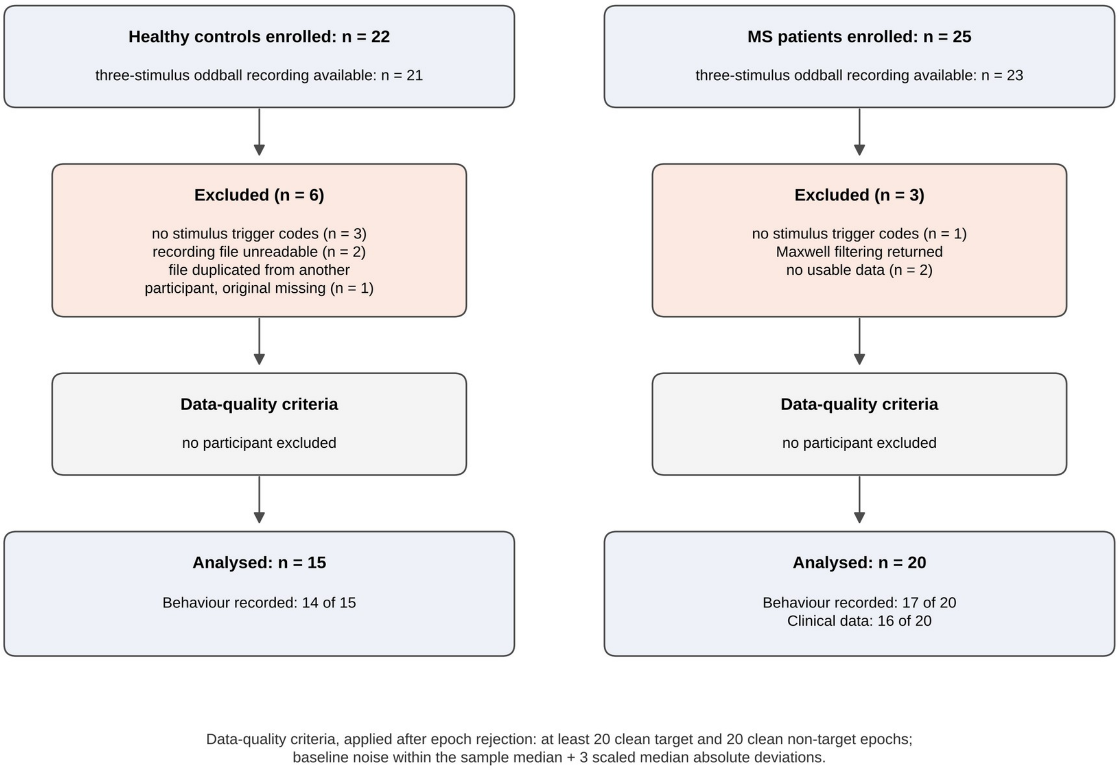


**Figure S1.** Participant flow. Participants enrolled, three-stimulus oddball recordings available on the analysis server, exclusions with reasons, and final sample. Data-quality criteria were applied after epoch rejection and excluded no participant.

**Figure S2**

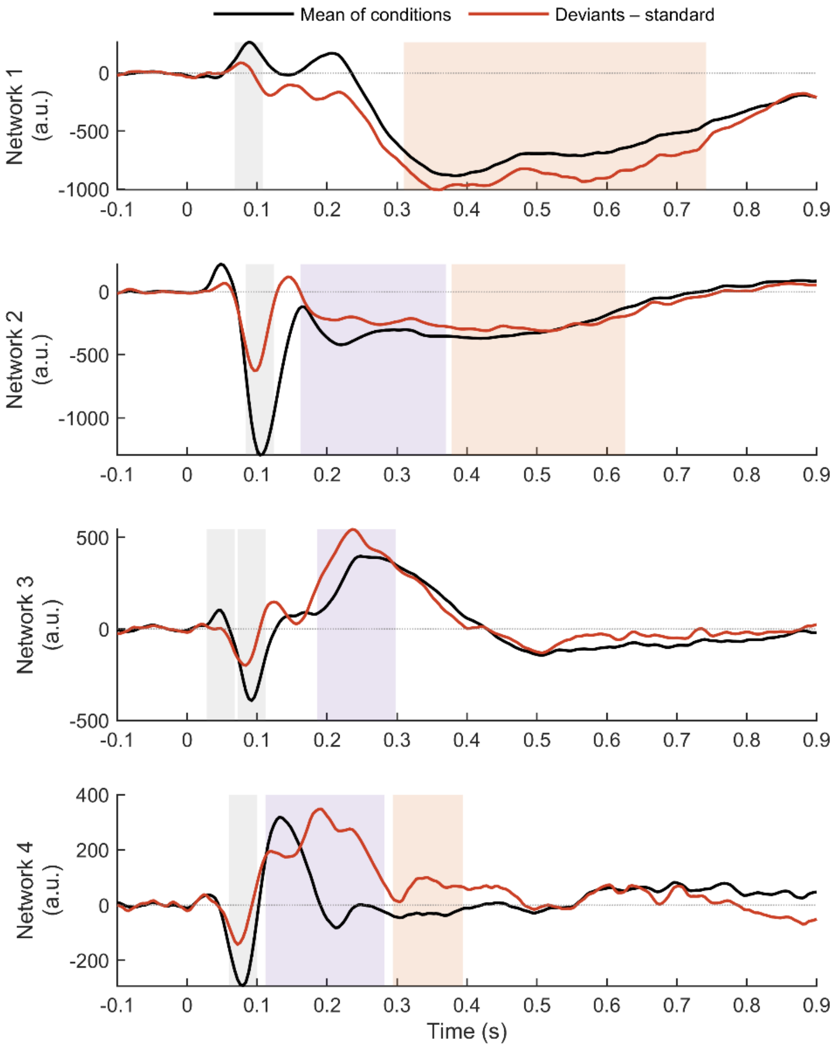


**Figure S2.** Localizer signals and analysis windows. For each network, the mean of the three conditions (black) and the deviant − standard difference (red), averaged with equal weight for the two groups and smoothed with a 20-ms moving average. Shaded areas mark the analysis windows, coloured by the signal that defined them (grey: mean of conditions; orange: deviant − standard; purple: both).

#### Figure S3

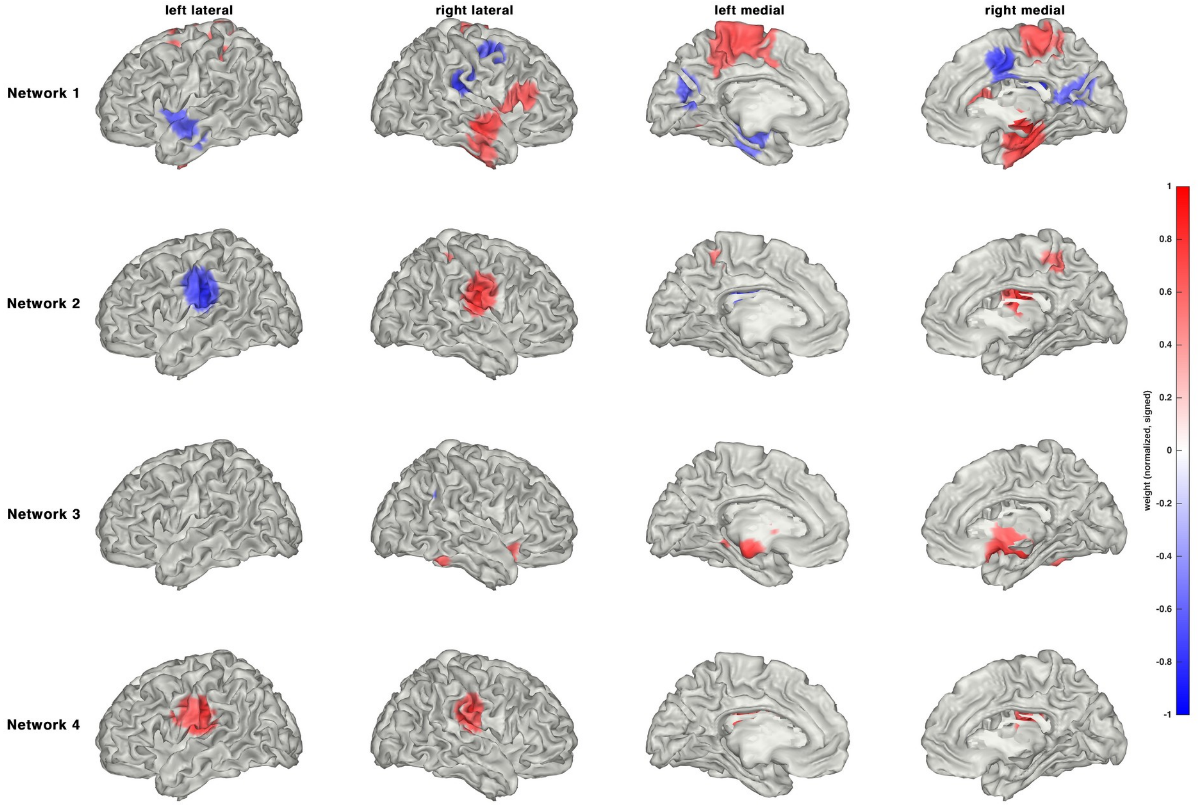


**Figure S3.** Signed spatial patterns of the four networks, projected on the cortical surface as in Fig. 2A (weights normalized to the maximum absolute weight of each network, shown above 50%). Red and blue indicate opposite signs; the overall sign of each network is arbitrary.

**Figure S4**

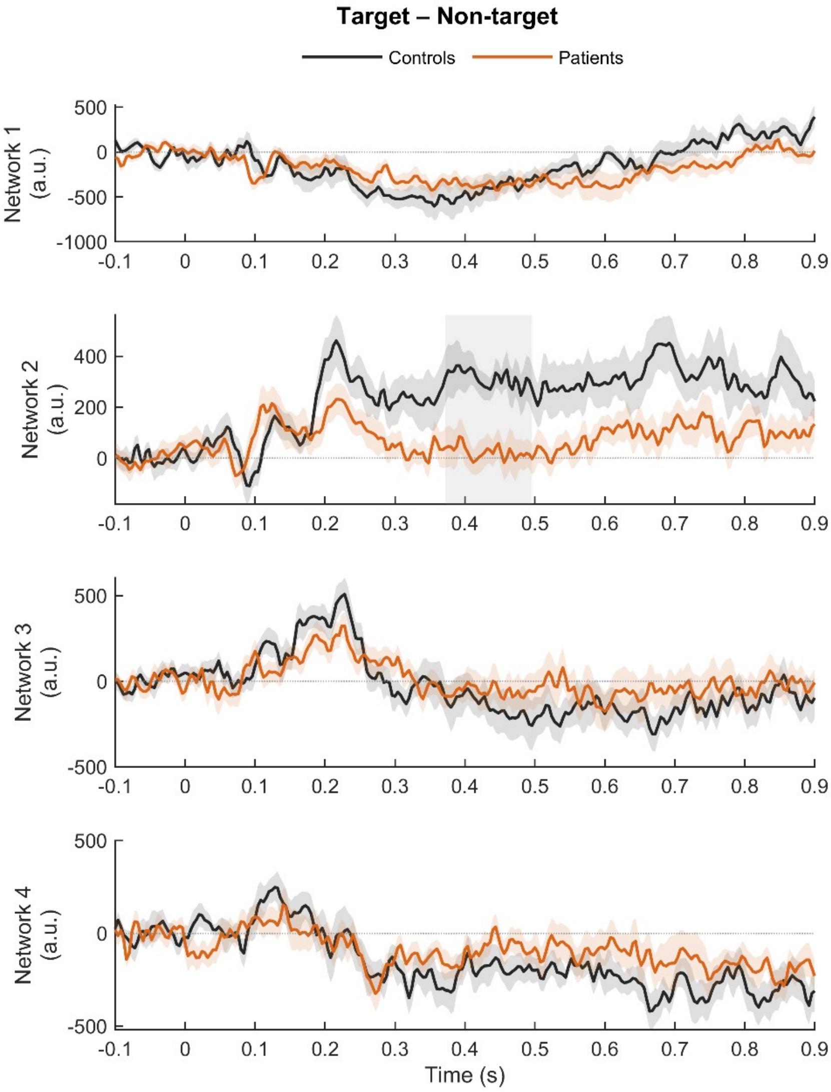


**Figure S4.** Target − non-target difference in the four networks for HC and MS patients (mean ± SEM). Grey bands mark clusters with a significant group difference after correction across networks (network 2 only).

#### Figure S5

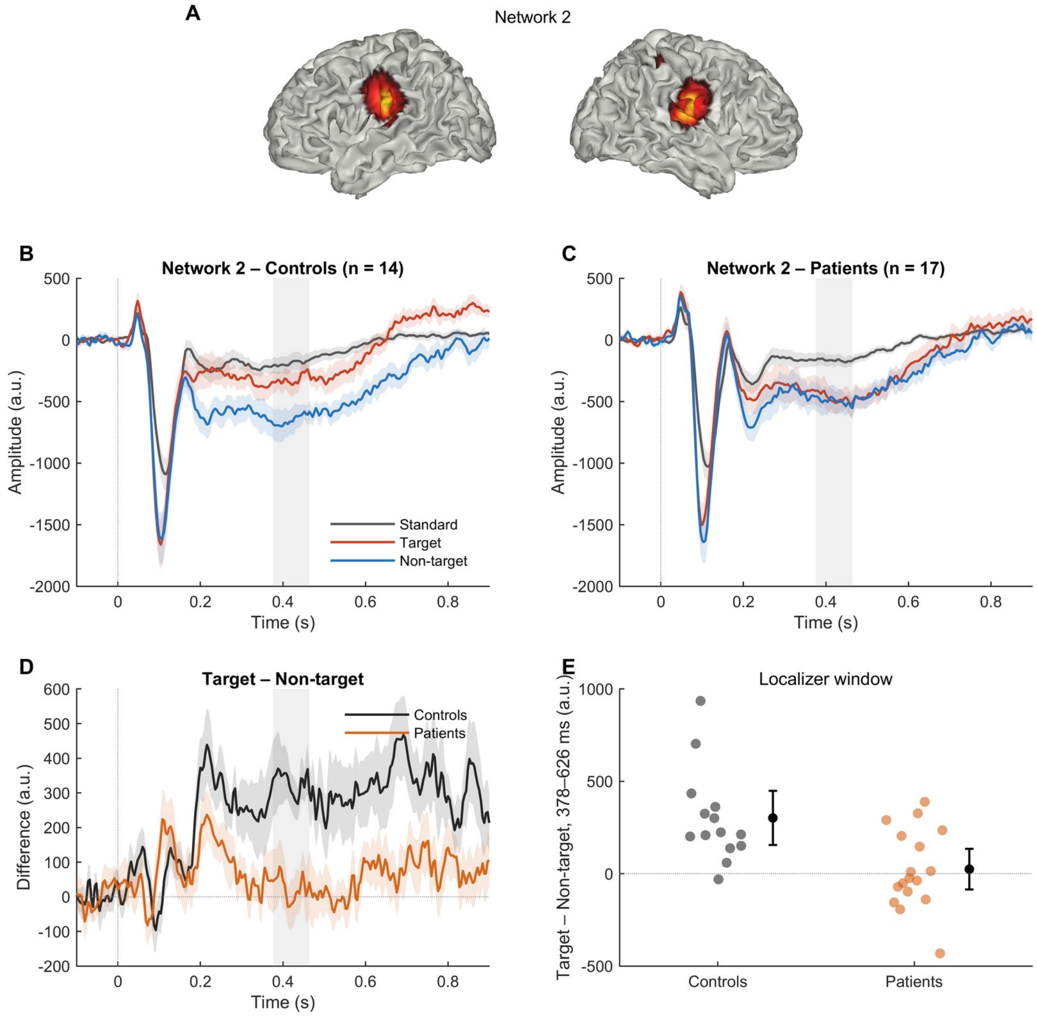


**Figure S5.** Sensitivity analysis: as Fig. 4, excluding four participants (14 HC, 17 MS patients; Section 2.5). The grey band marks the cluster at 376–464 ms ($p = 0.041$, corrected across networks). Localizer window: $g = -1.16$ [−1.92, −0.40], $p = 0.003$, BF10 = 14.4.

#### Figure S6

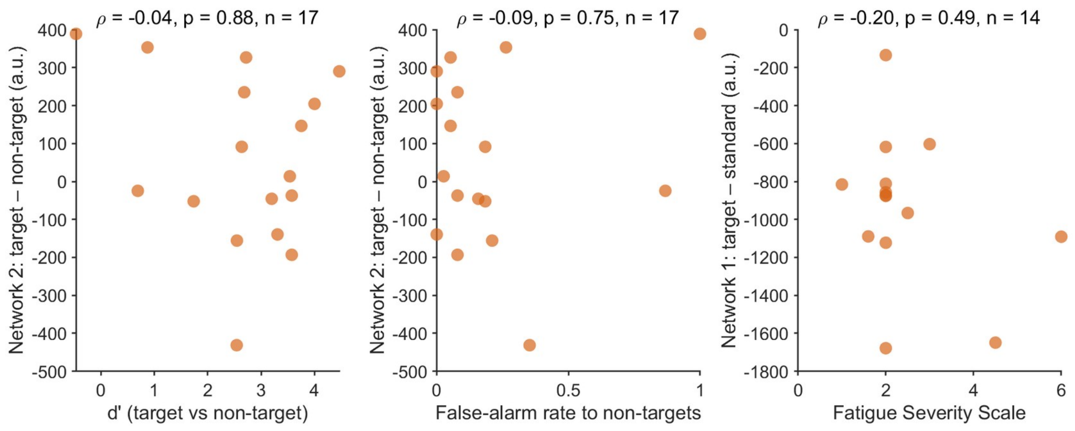


**Figure S6.** Relationships specified a priori in MS patients: network 2 target − non-target amplitude (378–626 ms) against d′ and against the false-alarm rate to non-targets (n = 17), and network 1 target − standard amplitude (310–742 ms) against the Fatigue Severity Scale (n = 14). Spearman correlations are reported in each panel.

#### Figure S7

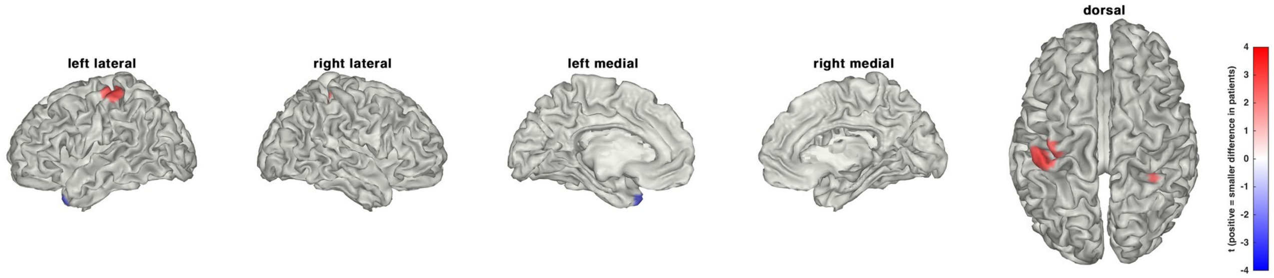


**Figure S7.** Source-level distribution of the group difference in the target − non-target amplitude (378–626 ms). The full map of Welch t values was projected on the cortical surface and is shown where |t| > 2.03 after projection; positive values indicate a smaller difference in MS patients. No spatial cluster survived correction across sources (Table S9); the map is descriptive.

### Supplementary tables

#### Table S1

Anatomical labels of the 5% of sources with the largest absolute weight in each network (AAL atlas; up to ten labels per network).

| Network | Region | Sources | Mean weight |
|---|---|---|---|
| 1 | Middle cingulate cortex (R) | 21 | −0.019 |
| 1 | Hippocampus (R) | 19 | +0.027 |
| 1 | Outside AAL labels | 16 | −0.027 |
| 1 | Supplementary motor area (R) | 13 | −0.007 |
| 1 | Precentral gyrus (R) | 12 | −0.026 |
| 1 | Superior temporal gyrus (R) | 10 | +0.011 |
| 1 | Parahippocampal gyrus (R) | 10 | +0.030 |
| 1 | Thalamus (R) | 8 | +0.015 |
| 1 | Middle cingulate cortex (L) | 7 | +0.020 |
| 1 | Supramarginal gyrus (R) | 6 | −0.032 |
| 2 | Postcentral gyrus (R) | 25 | +0.044 |
| 2 | Postcentral gyrus (L) | 21 | −0.048 |
| 2 | Rolandic operculum (R) | 14 | +0.058 |
| 2 | Supramarginal gyrus (L) | 13 | −0.045 |
| 2 | Insula (R) | 12 | +0.052 |
| 2 | Superior temporal gyrus (R) | 10 | +0.038 |
| 2 | Rolandic operculum (L) | 8 | −0.055 |
| 2 | Superior temporal gyrus (L) | 8 | −0.037 |
| 2 | Thalamus (R) | 7 | +0.046 |
| 2 | Insula (L) | 6 | −0.049 |
| 3 | Outside AAL labels | 40 | +0.043 |
| 3 | Caudate nucleus (R) | 12 | +0.041 |
| 3 | Putamen (R) | 11 | +0.026 |
| 3 | Thalamus (R) | 9 | +0.046 |
| 3 | Cerebellum, lobules IV–V (R) | 8 | +0.008 |
| 3 | Supramarginal gyrus (R) | 8 | −0.024 |
| 3 | Insula (R) | 7 | +0.005 |
| 3 | Parahippocampal gyrus (R) | 6 | +0.044 |
| 3 | Olfactory cortex (R) | 6 | +0.041 |
| 3 | Cerebellum, lobule VI (R) | 6 | −0.026 |
| 4 | Postcentral gyrus (L) | 17 | +0.046 |
| 4 | Postcentral gyrus (R) | 14 | +0.044 |
| 4 | Precentral gyrus (L) | 14 | +0.042 |
| 4 | Thalamus (L) | 13 | +0.038 |
| 4 | Outside AAL labels | 13 | +0.045 |
| 4 | Rolandic operculum (R) | 10 | +0.053 |
| 4 | Insula (R) | 10 | +0.050 |
| 4 | Rolandic operculum (L) | 10 | +0.051 |
| 4 | Insula (L) | 9 | +0.056 |
| 4 | Supramarginal gyrus (R) | 9 | +0.047 |

Each source took the most frequent AAL label within 6 mm of its position; Outside AAL labels: no label within 6 mm. Mean weight: mean signed weight of the sources with that label.

#### Table S2

Analysis windows defined by the collapsed localizer.

| Network | Window (ms) | Localizer signal |
|---|---|---|
| 1 | 68–108 | Mean of conditions |
| 1 | 310–742 | Deviants − standard |

| Network | Window (ms) | Localizer signal |
|---|---|---|
| 2 | 84–124 | Mean of conditions |
| 2 | 162–370 | Both |
| 2 | 378–626 | Deviants − standard |
| 3 | 28–68 | Mean of conditions |
| 3 | 72–112 | Mean of conditions |
| 3 | 186–298 | Both |
| 4 | 60–100 | Mean of conditions |
| 4 | 112–282 | Both |
| 4 | 294–394 | Deviants − standard |

Mean of conditions: peaks searched between 30 and 300 ms; deviants − standard: peaks searched between 150 and 700 ms; both: windows found in the two signals and merged.

#### Table S3

Window-based measures: condition effects and group comparisons.

| Network | Window (ms) | Measure | HC, mean (SD) | MS, mean (SD) | t (df) | p | Hedges' g [95% CI] | BF10 | Condition p |
|---|---|---|---|---|---|---|---|---|---|
| 1 | 68–108 | Standard | 202 (268) | 189 (206) | −0.16 (25.4) | 0.875 | −0.06 [−0.72, 0.61] | 0.33 | – |
| 1 | 68–108 | Target | 207 (470) | 120 (304) | −0.63 (22.5) | 0.535 | −0.22 [−0.89, 0.45] | 0.39 | – |
| 1 | 68–108 | Non-target | 215 (544) | 302 (258) | 0.57 (18.7) | 0.576 | 0.21 [−0.46, 0.88] | 0.38 | – |
| 1 | 68–108 | Target − standard | 6 (245) | −69 (337) | −0.76 (33.0) | 0.455 | −0.24 [−0.91, 0.43] | 0.40 | 0.470 |
| 1 | 68–108 | Non-target − standard | 13 (344) | 113 (219) | 0.98 (22.3) | 0.337 | 0.35 [−0.33, 1.02] | 0.50 | 0.147 |
| 1 | 68–108 | Target − non-target | −8 (393) | −182 (266) | −1.48 (23.2) | 0.153 | −0.52 [−1.20, 0.16] | 0.83 | 0.065 |
| 1 | 310–742 | Standard | −145 (124) | −103 (140) | 0.93 (32.0) | 0.361 | 0.30 [−0.37, 0.98] | 0.45 | – |
| 1 | 310–742 | Target | −1189 (439) | −1067 (441) | 0.81 (30.4) | 0.423 | 0.27 [−0.40, 0.94] | 0.42 | – |
| 1 | 310–742 | Non-target | −930 (362) | −756 (272) | 1.56 (25.1) | 0.130 | 0.54 [−0.14, 1.23] | 0.90 | – |
| 1 | 310–742 | Target − standard | −1045 (362) | −964 (441) | 0.59 (32.7) | 0.556 | 0.19 [−0.48, 0.86] | 0.37 | < 0.001 |
| 1 | 310–742 | Non-target − standard | −786 (304) | −653 (269) | 1.35 (28.1) | 0.189 | 0.46 [−0.22, 1.14] | 0.67 | < 0.001 |
| 1 | 310–742 | Target − non-target | −259 (356) | −312 (398) | −0.41 (31.9) | 0.685 | −0.13 [−0.80, 0.54] | 0.35 | < 0.001 |
| 2 | 84–124 | Standard | −886 (357) | −757 (455) | 0.94 (32.9) | 0.353 | 0.30 [−0.37, 0.98] | 0.45 | – |
| 2 | 84–124 | Target | −1351 (580) | −1150 (581) | 1.01 (30.3) | 0.320 | 0.34 [−0.34, 1.01] | 0.49 | – |
| 2 | 84–124 | Non-target | −1342 (620) | −1268 (599) | 0.36 (29.7) | 0.724 | 0.12 [−0.55, 0.79] | 0.34 | – |
| 2 | 84–124 | Target − standard | −465 (326) | −394 (356) | 0.61 (31.6) | 0.544 | 0.20 [−0.47, 0.87] | 0.38 | < 0.001 |
| 2 | 84–124 | Non-target − standard | −456 (367) | −511 (439) | −0.40 (32.5) | 0.690 | −0.13 [−0.80, 0.54] | 0.35 | < 0.001 |
| 2 | 84–124 | Target − non-target | −8 (155) | 117 (272) | 1.73 (31.1) | 0.094 | 0.53 [−0.15, 1.22] | 0.87 | 0.120 |
| 2 | 162–370 | Standard | −177 (235) | −175 (203) | 0.02 (27.7) | 0.984 | 0.01 [−0.66, 0.68] | 0.33 | – |
| 2 | 162–370 | Target | −278 (362) | −340 (415) | −0.48 (32.2) | 0.638 | −0.16 [−0.83, 0.51] | 0.36 | – |
| 2 | 162–370 | Non-target | −535 (383) | −435 (415) | 0.74 (31.5) | 0.462 | 0.25 [−0.43, 0.92] | 0.40 | – |
| 2 | 162–370 | Target − standard | −101 (202) | −165 (263) | −0.82 (33.0) | 0.421 | −0.26 [−0.93, 0.41] | 0.42 | 0.002 |
| 2 | 162–370 | Non-target − standard | −359 (336) | −259 (256) | 0.96 (25.3) | 0.348 | 0.33 [−0.34, 1.01] | 0.48 | < 0.001 |
| 2 | 162–370 | Target − non-target | 258 (234) | 94 (182) | −2.24 (25.6) | 0.034 | −0.78 [−1.47, −0.08] | 2.46 | < 0.001 |
| 2 | 378–626 | Standard | −118 (171) | −107 (136) | 0.20 (26.1) | 0.843 | 0.07 [−0.60, 0.74] | 0.33 | – |
| 2 | 378–626 | Target | −250 (272) | −357 (331) | −1.04 (32.7) | 0.304 | −0.34 [−1.01, 0.34] | 0.49 | – |
| 2 | 378–626 | Non-target | −547 (309) | −398 (317) | 1.40 (30.7) | 0.171 | 0.47 [−0.21, 1.14] | 0.69 | – |
| 2 | 378–626 | Target − standard | −132 (247) | −249 (288) | −1.29 (32.3) | 0.205 | −0.42 [−1.10, 0.25] | 0.61 | < 0.001 |
| 2 | 378–626 | Non-target − standard | −429 (208) | −290 (246) | 1.81 (32.5) | 0.080 | 0.59 [−0.10, 1.27] | 1.06 | < 0.001 |
| 2 | 378–626 | Target − non-target | 297 (245) | 41 (212) | −3.24 (27.6) | 0.003 | −1.11 [−1.82, −0.39] | 16.08 | 0.001 |
| 3 | 28–68 | Standard | 65 (136) | 49 (88) | −0.40 (22.7) | 0.696 | −0.14 [−0.81, 0.53] | 0.35 | – |
| 3 | 28–68 | Target | 25 (217) | 42 (178) | 0.24 (26.7) | 0.814 | 0.08 [−0.59, 0.75] | 0.34 | – |

| Network | Window (ms) | Measure | HC, mean (SD) | MS, mean (SD) | t (df) | p | Hedges' g [95% CI] | BF10 | Condition p |
|---|---|---|---|---|---|---|---|---|---|
| 3 | 28–68 | Non-target | −25 (158) | 95 (184) | 2.07 (32.3) | 0.046 | 0.68 [−0.01, 1.36] | 1.54 | – |
| 3 | 28–68 | Target − standard | −39 (155) | −7 (177) | 0.57 (32.1) | 0.570 | 0.19 [−0.48, 0.86] | 0.37 | 0.462 |
| 3 | 28–68 | Non-target − standard | −90 (167) | 46 (180) | 2.31 (31.5) | 0.027 | 0.76 [0.07, 1.46] | 2.30 | 0.704 |
| 3 | 28–68 | Target − non-target | 50 (168) | −53 (287) | −1.34 (31.4) | 0.190 | −0.42 [−1.09, 0.26] | 0.60 | 0.831 |
| 3 | 72–112 | Standard | −275 (216) | −168 (270) | 1.31 (32.8) | 0.200 | 0.42 [−0.25, 1.10] | 0.61 | – |
| 3 | 72–112 | Target | −405 (438) | −184 (339) | 1.62 (25.6) | 0.117 | 0.56 [−0.12, 1.24] | 0.96 | – |
| 3 | 72–112 | Non-target | −464 (446) | −243 (366) | 1.57 (26.7) | 0.129 | 0.54 [−0.14, 1.22] | 0.88 | – |
| 3 | 72–112 | Target − standard | −130 (291) | −17 (218) | 1.26 (25.0) | 0.218 | 0.44 [−0.24, 1.12] | 0.64 | 0.139 |
| 3 | 72–112 | Non-target − standard | −189 (308) | −75 (185) | 1.27 (21.4) | 0.218 | 0.45 [−0.22, 1.13] | 0.67 | 0.006 |
| 3 | 72–112 | Target − non-target | 60 (188) | 59 (199) | −0.01 (31.2) | 0.991 | −0.00 [−0.67, 0.67] | 0.33 | 0.077 |
| 3 | 186–298 | Standard | 28 (202) | 3 (173) | −0.39 (27.4) | 0.703 | −0.13 [−0.80, 0.54] | 0.35 | – |
| 3 | 186–298 | Target | 606 (461) | 496 (461) | −0.70 (30.3) | 0.491 | −0.23 [−0.90, 0.44] | 0.40 | – |
| 3 | 186–298 | Non-target | 370 (282) | 315 (330) | −0.53 (32.4) | 0.598 | −0.17 [−0.84, 0.50] | 0.36 | – |
| 3 | 186–298 | Target − standard | 578 (406) | 493 (380) | −0.63 (29.2) | 0.535 | −0.21 [−0.88, 0.46] | 0.38 | < 0.001 |
| 3 | 186–298 | Non-target − standard | 342 (226) | 312 (235) | −0.38 (30.9) | 0.704 | −0.13 [−0.80, 0.54] | 0.35 | < 0.001 |
| 3 | 186–298 | Target − non-target | 236 (241) | 182 (264) | −0.64 (31.7) | 0.529 | −0.21 [−0.88, 0.46] | 0.38 | < 0.001 |
| 4 | 60–100 | Standard | −199 (263) | −147 (267) | 0.58 (30.6) | 0.568 | 0.19 [−0.48, 0.86] | 0.37 | – |
| 4 | 60–100 | Target | −257 (426) | −223 (385) | 0.24 (28.5) | 0.810 | 0.08 [−0.59, 0.75] | 0.34 | – |
| 4 | 60–100 | Non-target | −249 (429) | −256 (369) | −0.05 (27.6) | 0.960 | −0.02 [−0.69, 0.65] | 0.33 | – |
| 4 | 60–100 | Target − standard | −58 (234) | −76 (262) | −0.22 (31.9) | 0.829 | −0.07 [−0.74, 0.60] | 0.33 | 0.110 |
| 4 | 60–100 | Non-target − standard | −50 (217) | −109 (223) | −0.79 (30.7) | 0.437 | −0.26 [−0.93, 0.41] | 0.42 | 0.030 |
| 4 | 60–100 | Target − non-target | −8 (196) | 33 (213) | 0.59 (31.6) | 0.560 | 0.19 [−0.48, 0.86] | 0.37 | 0.659 |
| 4 | 112–282 | Standard | −82 (271) | −59 (203) | 0.28 (24.9) | 0.785 | 0.10 [−0.57, 0.77] | 0.34 | – |
| 4 | 112–282 | Target | 196 (403) | 117 (386) | −0.59 (29.6) | 0.561 | −0.20 [−0.87, 0.47] | 0.38 | – |
| 4 | 112–282 | Non-target | 179 (320) | 157 (313) | −0.21 (29.9) | 0.839 | −0.07 [−0.74, 0.60] | 0.33 | – |
| 4 | 112–282 | Target − standard | 278 (345) | 176 (300) | −0.92 (27.8) | 0.366 | −0.31 [−0.99, 0.36] | 0.46 | < 0.001 |
| 4 | 112–282 | Non-target − standard | 262 (254) | 216 (242) | −0.53 (29.5) | 0.599 | −0.18 [−0.85, 0.49] | 0.37 | < 0.001 |
| 4 | 112–282 | Target − non-target | 17 (277) | −40 (232) | −0.65 (27.1) | 0.522 | −0.22 [−0.89, 0.45] | 0.39 | 0.711 |
| 4 | 294–394 | Standard | −109 (130) | −46 (147) | 1.35 (32.0) | 0.186 | 0.44 [−0.23, 1.12] | 0.64 | – |
| 4 | 294–394 | Target | −126 (385) | −86 (287) | 0.34 (24.9) | 0.740 | 0.12 [−0.55, 0.79] | 0.34 | – |
| 4 | 294–394 | Non-target | 120 (348) | 51 (360) | −0.57 (30.9) | 0.576 | −0.19 [−0.86, 0.48] | 0.37 | – |
| 4 | 294–394 | Target − standard | −17 (434) | −40 (309) | −0.18 (24.1) | 0.857 | −0.06 [−0.73, 0.61] | 0.33 | 0.624 |
| 4 | 294–394 | Non-target − standard | 229 (389) | 97 (389) | −0.99 (30.3) | 0.329 | −0.33 [−1.01, 0.34] | 0.48 | 0.026 |
| 4 | 294–394 | Target − non-target | −245 (294) | −137 (182) | 1.25 (21.9) | 0.223 | 0.45 [−0.23, 1.12] | 0.65 | < 0.001 |

Amplitudes in a.u. t (df): Welch's t-test, MS patients vs HC. g: Hedges' g (MS − HC). BF10: Jeffreys–Zellner–Siow Bayes factor (Cauchy prior scale √2/2). Condition p: one-sample t-test of the contrast across all participants (difference measures only). All p values uncorrected.

#### Table S4

Latencies (50% fractional area, jackknife).

| Network | Window (ms) | Condition | HC (ms) | MS (ms) | Difference (ms) | t | p |
|---|---|---|---|---|---|---|---|
| 1 | 68–108 | Standard | 91 | 90 | −1 | −0.39 | 0.697 |
| 1 | 68–108 | Target | 85 | 80 | −5 | −1.44 | 0.160 |
| 1 | 68–108 | Non-target | 87 | 88 | 1 | 0.20 | 0.840 |
| 1 | 310–742 | Target | 487 | 502 | 16 | 1.55 | 0.131 |
| 1 | 310–742 | Non-target | 517 | 503 | −13 | −1.45 | 0.156 |
| 2 | 84–124 | Standard | 105 | 106 | 0 | 0.29 | 0.770 |
| 2 | 84–124 | Target | 102 | 101 | −1 | −0.43 | 0.673 |
| 2 | 84–124 | Non-target | 103 | 103 | −0 | −0.36 | 0.722 |
| 2 | 162–370 | Standard | 266 | 242 | −24 | −1.30 | 0.201 |
| 2 | 162–370 | Target | 278 | 267 | −11 | −0.75 | 0.458 |
| 2 | 162–370 | Non-target | 267 | 255 | −11 | −1.07 | 0.292 |
| 2 | 378–626 | Target | 481 | 476 | −5 | −0.25 | 0.805 |
| 2 | 378–626 | Non-target | 488 | 482 | −5 | −0.81 | 0.426 |
| 3 | 28–68 | Standard | 45 | 44 | −1 | −0.26 | 0.793 |
| 3 | 28–68 | Target | 44 | 42 | −2 | −0.61 | 0.547 |
| 3 | 28–68 | Non-target | 44 | 44 | 0 | 0.01 | 0.994 |
| 3 | 72–112 | Standard | 94 | 95 | 1 | 0.34 | 0.737 |
| 3 | 72–112 | Target | 88 | 86 | −2 | −0.68 | 0.500 |
| 3 | 72–112 | Non-target | 90 | 90 | −1 | −0.21 | 0.837 |
| 3 | 186–298 | Standard | 267 | 275 | 8 | 1.43 | 0.163 |
| 3 | 186–298 | Target | 239 | 247 | 8 | 1.36 | 0.184 |
| 3 | 186–298 | Non-target | 256 | 257 | 1 | 0.19 | 0.847 |
| 4 | 60–100 | Standard | 81 | 77 | −4 | −0.60 | 0.552 |
| 4 | 60–100 | Target | 74 | 76 | 1 | 0.27 | 0.791 |
| 4 | 60–100 | Non-target | 74 | 78 | 3 | 0.61 | 0.545 |
| 4 | 112–282 | Standard | 134 | 136 | 2 | 0.27 | 0.787 |
| 4 | 112–282 | Target | 143 | 145 | 2 | 0.11 | 0.914 |
| 4 | 112–282 | Non-target | 157 | 163 | 6 | 0.24 | 0.808 |
| 4 | 294–394 | Target | – | – | – | – | – |
| 4 | 294–394 | Non-target | 345 | 363 | 18 | 0.61 | 0.548 |

Difference: MS − HC. t: jackknife-corrected t-statistic (df = 33). –: latency not computable (no area of the expected polarity in the window). For windows defined only on the deviant − standard signal, latencies were computed for target and non-target responses only.

#### Table S5

Data quality by group.

| Measure | HC | MS | p |
|---|---|---|---|
| Retained standard epochs, mean (range) | 293.1 (190–304) | 298.4 (260–304) | 0.53 |
| Retained target epochs, mean (range) | 36.8 (23–38) | 36.2 (20–38) | 0.67 |
| Retained non-target epochs, mean (range) | 37.1 (26–38) | 36.7 (29–38) | 0.72 |
| ICA components removed, mean ± SD (range) | 2.6 ± 0.9 (2–5) | 3.4 ± 1.5 (2–7) | 0.08 |
| Sensor baseline noise, fT, median (range) | 159 (114–282) | 180 (123–257) | 0.418 |
| N1m signal-to-noise ratio (network 2, standard), median | 31.0 | 25.6 | 0.97 |
| Sources with inverted sign, %, mean ± SD | 33.1 ± 4.2 | 31.9 ± 4.5 | 0.418 |

p: Welch's t-test, MS patients vs HC (medians are shown for skewed measures). ICA components counted from the reduction in data rank.

Baseline noise of the network time series (median across participants of the root mean square from −100 to 0 ms, a.u.; p: Welch's t-test).

| Network | Condition | HC | MS | p |
|---|---|---|---|---|
| 1 | Standard | 59.3 | 50.8 | 0.456 |
| 1 | Target | 146.3 | 163.5 | 0.902 |

| Network | Condition | HC | MS | p |
|---|---|---|---|---|
| 1 | Non-target | 146.1 | 155.0 | 0.734 |
| 2 | Standard | 36.9 | 40.8 | 0.801 |
| 2 | Target | 98.6 | 109.2 | 0.803 |
| 2 | Non-target | 89.1 | 89.7 | 0.665 |
| 3 | Standard | 47.0 | 45.7 | 0.771 |
| 3 | Target | 128.5 | 149.8 | 0.262 |
| 3 | Non-target | 129.6 | 125.3 | 0.689 |
| 4 | Standard | 44.4 | 54.8 | 0.466 |
| 4 | Target | 125.8 | 142.8 | 0.197 |
| 4 | Non-target | 123.2 | 142.1 | 0.595 |

#### Table S6

Robustness of the main result (network 2, target − non-target).

| Analysis | Result |
|---|---|
| Main analysis (15 HC, 20 MS) | Localizer window 378–626 ms: g = −1.11 [−1.82, −0.39], p = 0.003, BF10 = 16.1; cluster 372–496 ms, p = 0.010 (network 2), p = 0.020 (corrected across networks) |
| Correction across all contrasts and networks | Same cluster: p = 0.108 |
| Leave-one-participant-out (localizer window) | g between −1.23 and −1.02; p between 0.002 and 0.006 |
| Networks estimated with equal group weights | \|r\| ≥ 0.99 with the original networks; g = −1.10, p = 0.003; cluster 372–496 ms, p = 0.006 (network 2) |
| Sensitivity sample (14 HC, 17 MS) | g = −1.16 [−1.92, −0.40], p = 0.003, BF10 = 14.4; cluster 376–464 ms, p = 0.013 (network 2), p = 0.041 (corrected across networks) |
| Excluding participants who also responded to non-targets (14 HC, 18 MS) | g = −1.22 [−1.97, −0.46], p = 0.002, BF10 = 22.5; cluster 368–500 ms, p = 0.006 (network 2), p = 0.016 (corrected across networks) |
| ICA components removed as covariate | Group effect −233 a.u., t = −2.89, p = 0.007; components: t = −1.03, p = 0.31; Spearman ρ with the effect = −0.15, p = 0.39 |
| Source-level analysis (3559 sources) | No spatial cluster significant after correction (largest cluster p = 0.35) |

#### Table S7

Correlations with behaviour and clinical measures in MS patients (Spearman).

| Type | Network measure | Variable | ρ | p | q (FDR) | n |
|---|---|---|---|---|---|---|
| A priori | Network 2, target − non-target | d′ | −0.04 | 0.885 | – | 17 |
| A priori | Network 2, target − non-target | False-alarm rate (non-targets) | −0.09 | 0.745 | – | 17 |
| A priori | Network 1, target − standard | FSS | −0.20 | 0.494 | – | 14 |
| Exploratory | Network 2, target − non-target | SDMT | −0.54 | 0.030 | 0.32 | 16 |
| Exploratory | Network 2, target − non-target | Number of altered tests | +0.54 | 0.032 | 0.32 | 16 |
| Exploratory | Network 1, target − standard | SDMT | −0.45 | 0.082 | 0.42 | 16 |
| Exploratory | Network 1, target − standard | Hit rate | −0.42 | 0.092 | 0.42 | 17 |
| Exploratory | Network 2, target − non-target | Total lesion count | +0.42 | 0.108 | 0.42 | 16 |
| Exploratory | Network 1, target − standard | Disease duration | +0.40 | 0.126 | 0.42 | 16 |
| Exploratory | Network 1, target − standard | BDI-II | +0.38 | 0.152 | 0.44 | 16 |
| Exploratory | Network 2, target − non-target | Hit rate | −0.30 | 0.238 | 0.51 | 17 |
| Exploratory | Network 1, target − standard | Number of altered tests | +0.30 | 0.251 | 0.51 | 16 |
| Exploratory | Network 1, target − standard | Median reaction time | +0.28 | 0.276 | 0.51 | 17 |
| Exploratory | Network 2, target − non-target | BDI-II | +0.28 | 0.291 | 0.51 | 16 |
| Exploratory | Network 1, target − standard | d′ | −0.26 | 0.304 | 0.51 | 17 |
| Exploratory | Network 1, target − standard | Total lesion count | +0.26 | 0.337 | 0.52 | 16 |
| Exploratory | Network 2, target − non-target | Disease duration | +0.24 | 0.379 | 0.54 | 16 |
| Exploratory | Network 1, target − standard | PASAT-3 | −0.19 | 0.540 | 0.68 | 13 |
| Exploratory | Network 1, target − standard | EDSS | +0.16 | 0.576 | 0.68 | 15 |
| Exploratory | Network 2, target − non-target | PASAT-3 | +0.17 | 0.577 | 0.68 | 13 |

| Type | Network measure | Variable | ρ | p | q (FDR) | n |
|---|---|---|---|---|---|---|
| Exploratory | Network 2, target − non-target | FSS | +0.08 | 0.798 | 0.89 | 14 |
| Exploratory | Network 2, target − non-target | Median reaction time | +0.04 | 0.883 | 0.90 | 17 |
| Exploratory | Network 2, target − non-target | EDSS | −0.03 | 0.904 | 0.90 | 15 |

Network 2 measure: localizer window 378–626 ms; network 1 measure: 310–742 ms. q: Benjamini–Hochberg false discovery rate across the exploratory correlations.

#### Table S8

Neuropsychological scores by test in HC and MS patients.

| Test | HC, mean (SD) | n | MS, mean (SD) | n | Altered, HC · MS | p |
|---|---|---|---|---|---|---|
| SRT-LTS | 43.5 (13.0) | 13 | 37.4 (10.3) | 16 | 1/13 · 1/16 | 0.068 |
| SRT-CLTR | 32.2 (14.5) | 13 | 26.7 (11.7) | 16 | 1/13 · 1/16 | 0.174 |
| SPART | 20.2 (5.9) | 13 | 19.2 (6.5) | 16 | 1/13 · 1/16 | 0.709 |
| SDMT | 53.2 (13.7) | 13 | 47.2 (13.9) | 16 | 0/13 · 2/16 | 0.272 |
| PASAT-3 | 42.6 (8.8) | 12 | 31.2 (12.9) | 13 | 1/13 · 4/16 | 0.022 |
| PASAT-5 | 50.3 (6.0) | 12 | 44.0 (9.4) | 13 | 1/13 · 3/16 | 0.067 |
| SRT-D | 7.9 (3.1) | 13 | 7.6 (2.2) | 16 | 1/13 · 0/16 | 0.340 |
| SPART-D | 6.7 (2.9) | 13 | 6.6 (2.7) | 16 | 1/13 · 3/16 | 0.859 |
| WLG | 20.8 (5.3) | 13 | 15.7 (5.1) | 16 | 3/13 · 4/16 | 0.018 |
| TMT-A (s) | 30.2 (9.3) | 13 | 39.5 (14.6) | 15 | 0/13 · 0/15 | 0.111 |
| TMT-B (s) | 50.1 (22.4) | 13 | 73.6 (73.3) | 15 | 0/13 · 1/15 | 0.298 |

Raw scores of Rao's Brief Repeatable Battery and of the Trail Making Test (completion time in seconds); clinical data were available for 16 of the 20 patients and neuropsychological data for 13 of the 15 HC. PASAT scores of zero were treated as missing (one HC and three patients); the clinical team scored these tests as altered, and they are counted as such in the "Altered" column (number of participants with an altered test / number assessed). p: Mann–Whitney U test, uncorrected. SRT-LTS, long-term storage; SRT-CLTR, consistent long-term retrieval; SPART, 10/36 Spatial Recall Test; SRT-D and SPART-D, delayed recall; WLG, word list generation.

#### Table S9

Source-level clusters of the group difference in target − non-target (378–626 ms), clusters of at least 10 sources.

| Mass | Sources | p (corrected) | Peak MNI (mm) | Peak t | Peak g | AAL regions (sources) |
|---|---|---|---|---|---|---|
| 109.2 | 43 | 0.345 | −46, −30, 56 | 3.87 | 1.39 | Postcentral gyrus (L) 15; Middle temporal gyrus (L) 13; Precentral gyrus (L) 6; Inferior parietal lobule (L) 5; Supramarginal gyrus (L) 2; Angular gyrus (L) 1 |
| 77.0 | 31 | 0.534 | 26, −46, 56 | 3.11 | 1.02 | Postcentral gyrus (R) 9; Middle cingulate cortex (R) 7; Inferior parietal lobule (R) 6; Paracentral lobule (R) 2; Superior parietal lobule (R) 2; Angular gyrus (R) 1 |
| −42.4 | 17 | 0.840 | 10, −38, 40 | −3.33 | −1.09 | Precuneus (R) 6; Middle cingulate cortex (R) 5; Paracentral lobule (R) 4; Angular gyrus (R) 1; Superior parietal lobule (R) 1 |
| −31.5 | 13 | 0.928 | −22, 18, −40 | −2.79 | −0.88 | Temporal pole (middle) (L) 7; Temporal pole (superior) (L) 3; Inferior frontal gyrus (orbital) (L) 1; Gyrus rectus (L) 1; Inferior temporal gyrus (L) 1 |
| 31.4 | 14 | 0.929 | −46, 2, 32 | 2.56 | 0.87 | Precentral gyrus (L) 11; Postcentral gyrus (L) 2; Inferior frontal gyrus (opercular) (L) 1 |
| 28.1 | 12 | 0.954 | −14, −70, 0 | 3.03 | 0.98 | Lingual gyrus (L) 3; Calcarine cortex (L) 2; Cuneus (L) 2; Vermis, lobules IV–V 2; Calcarine cortex (R) 1; Cuneus (R) 1 |
| 24.6 | 10 | 0.973 | −14, 34, 24 | 3.26 | 1.08 | Middle frontal gyrus (L) 4; Superior frontal gyrus (L) 3; Anterior cingulate cortex (L) 2; Superior frontal gyrus (medial) (L) 1 |

Positive mass, t and g: smaller target − non-target difference in MS patients (each source oriented by the mean response of the two groups). p corrected for the family-wise error across all sources (5000 permutations). Regions: AAL labels of the cluster sources, with number of sources.
